# Clinical Relevance of Computationally Derived Tubular Features: Spatial Relationships and the Development of Tubulointerstitial Scarring in MCD/FSGS

**DOI:** 10.1101/2024.07.19.24310619

**Authors:** Fan Fan, Qian Liu, Jarcy Zee, Takaya Ozeki, Dawit Demeke, Yingbao Yang, Alton B. Farris, Bangcheng Wang, Manav Shah, Jackson Jacobs, Laura Mariani, Kyle Lafata, Jeremy Rubin, Yijiang Chen, Lawrence Holzman, Jeffrey B. Hodgin, Anant Madabhushi, Laura Barisoni, Andrew Janowczyk

## Abstract

**Background:** Visual scoring of tubular damage has limitations in capturing the full spectrum of structural changes and prognostic potential. We investigate if computationally quantified tubular features can enhance prognostication and reveal spatial relationships with interstitial fibrosis.

**Methods:** Deep-learning and image-processing-based segmentations were employed in N=254/266 PAS-WSIs from the NEPTUNE/CureGN datasets (135/153 focal segmental glomerulosclerosis and 119/113 minimal change disease) for: cortex, tubular lumen (TL), epithelium (TE), nuclei (TN), and basement membrane (TBM). N=104 pathomic features were extracted from these segmented tubular substructures and summarized at the patient level using summary statistics. The tubular features were quantified across the biopsy and in manually segmented regions of mature interstitial fibrosis and tubular atrophy (IFTA), pre-IFTA and non-IFTA in the NEPTUNE dataset. Minimum Redundancy Maximum Relevance was used in the NEPTUNE dataset to select features most associated with disease progression and proteinuria remission. Ridge-penalized Cox models evaluated their predictive discrimination compared to clinical/demographic data and visual-assessment. Models were evaluated in the CureGN dataset.

**Results:** N=9 features were predictive of disease progression and/or proteinuria remission. Models with tubular features had high prognostic accuracy in both NEPTUNE and CureGN datasets and increased prognostic accuracy for both outcomes (5.6%-7.7% and 1.6%-4.6% increase for disease progression and proteinuria remission, respectively) compared to conventional parameters alone in the NEPTUNE dataset. TBM thickness/area and TE simplification progressively increased from non- to pre- and mature IFTA.

**Conclusions:** Previously under-recognized, quantifiable, and clinically relevant tubular features in the kidney parenchyma can enhance understanding of mechanisms of disease progression and risk stratification.

## INTRODUCTION

Conventional assessment of the tubulointerstitial in kidney biopsies is based on visual scoring of a limited number of histologic parameters. These include continuous percentage or categorical scoring of interstitial fibrosis and tubular atrophy (IFTA) separately or combined as IFTA, interstitial inflammation, and acute tubular injury^1–7^. Although the association between these parameters with kidney function and disease progression has been shown by multiple studies in native and transplant kidney biopsies, their potential for predicting outcomes is limited due to known inter- and intra-observer variability and lack of standardized scoring systems^3,8,9^. Furthermore, encoded in the kidney tissue may be quantitative and topological information that the human vision system is not equipped to capture or quantify. These characteristics may have the potential to improve our ability to better characterize changes of individual functional tissue units, their spatial relationship, and to identify novel biopsy-based biomarkers of disease progression^1,10,11^. Thus, the precise and reproducible quantification of tubulointerstitial histologic characteristics can contribute to the understanding of structural and functional changes in kidney diseases, enhance prognostication and prediction of clinical outcome, and ultimately of clinical care^5,7,12^.

Digital and computational pathology has enabled precise tissue quantification through deep learning (DL) models that segment normal and abnormal functional tissue units, such as glomeruli, tubules, arteries, and peritubular capillaries^13–15^. Pathomic features, which are quantitative attributes from pathology images, capture morphological and structural characteristics of kidney structures^1,15,16^. These features represent the heterogeneity of kidney structural changes and can be extracted from segmented functional tissue units. They are essential for quantifying normal and pathological states, enabling precise tissue characterization, and enhancing our ability to diagnose and prognosticate kidney diseases in a reliable and reproducible manner^1^.

Leveraging previously established rich clinical and whole slide image (WSI) cohorts, such as those from the Nephrotic Syndrome Study Network (NEPTUNE)^17^ and Cure Glomerulonephropathy (CureGN)^18^ consortia, this study employs computational pathology. It tests the hypothesis that sophisticated and computationally assessed tubular features extracted from digital kidney biopsies of patients with focal segmental glomerulosclerosis (FSGS) and minimal change disease (MCD) can enhance the prognostication of glomerular diseases and define the progression from normal to severe kidney scarring. To test this hypothesis, we:

a. developed and applied multiple DL-based or traditional image-processing-based algorithms for the segmentation of different tubular substructures, including tubular basement membrane (TBM), tubular lumen (TL), tubular epithelium (TE), and tubular nuclei (TN);
b. used pathomic features to extract and quantify the morphometric characteristics of the tubular substructures;
c. identified the top features most predictive of clinical outcome and compared the prognostic value of these features to conventional pathology and clinical parameters;
d. studied the tubular features in pre-, mature, and non-IFTA regions and compared them to non-IFTA regions of reference tissue.

## METHODS

### 1. Study cohorts and sample collection

Three cohorts were used in this study: 1) NEPTUNE (n=254)^17^, training and internal validation cohort; 2) CureGN (n=266)^18^, external validation cohort; 3) Nephrectomies (n=13) from the University of Michigan (UMICH), reference tissue. Written informed consent was obtained from all NEPTUNE/CureGN participants and UMICH nephrectomy patients (**Figure 1**). A Periodic Acid Schiff (PAS) stained whole slide images (WSI) per participant was used.

**Figure 1:**
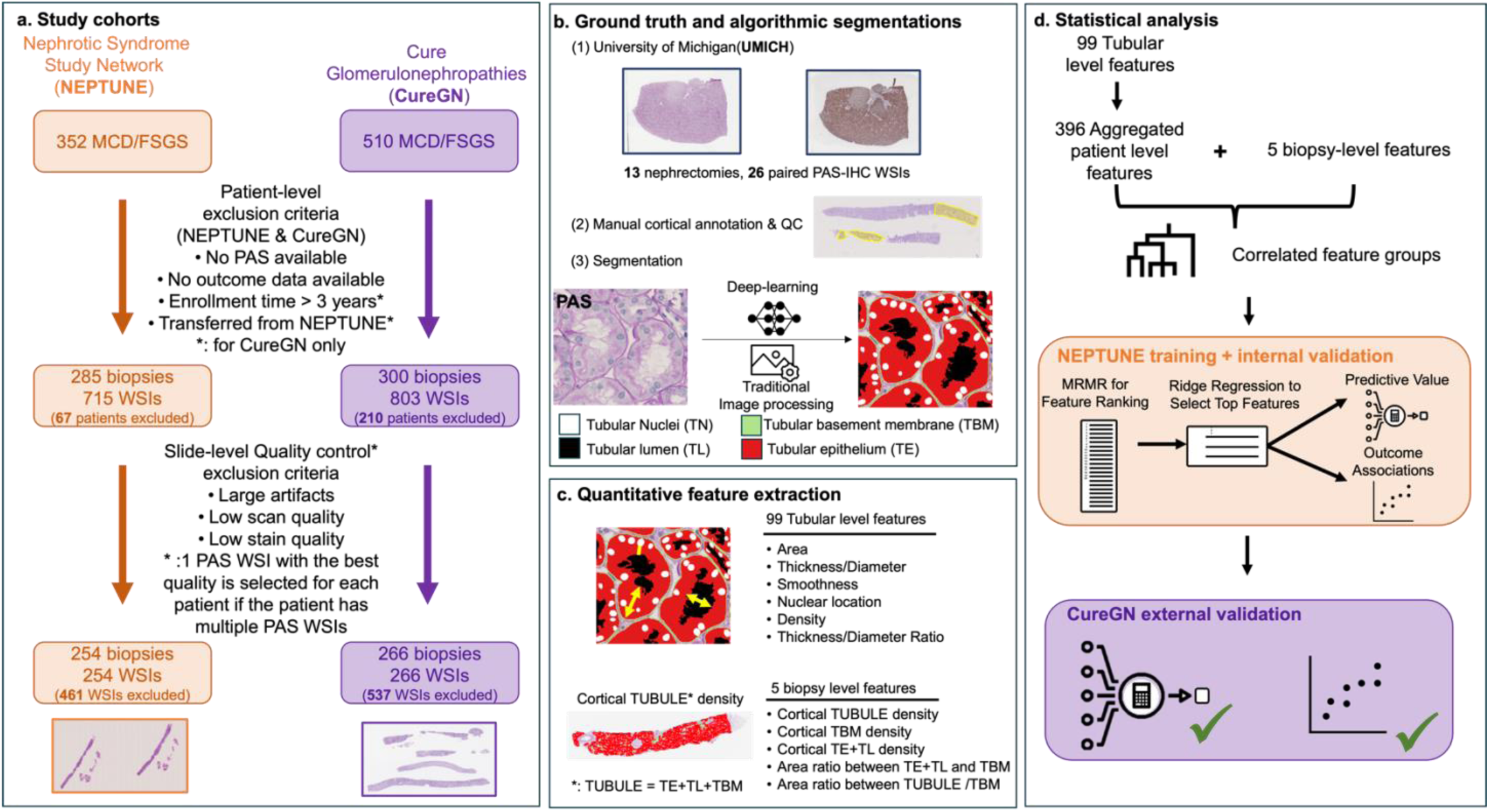
Overview of the workflow: **a.** Study cohorts: Depicts the study sample selection process, where all the patients and whole slide images (WSIs) were passed through each of the sample selection processes following the patient-level and slide-level exclusion criteria resulting in the inclusion of 520 patients from 2 cohorts and 520 WSIs in this study. **b.** Ground truth and algorithmic segmentations: (1) Reference tissue from the University of Michigan, (2) Cortical area was annotated and quality controlled (QCed) by study pathologists and (3) Utilization of both deep-learning (DL) and traditional image-processing algorithms to segment tubular primitives from the PAS WSIs. **c.** Quantitative feature extraction: Extraction of 99 tubule-level and 5 biopsy-level features from the segmentation results in **b** using image-processing algorithms. **d.** Statistical analysis: tubular level features were aggregated to patient-level features which, along with biopsy-level features, were being clustered into feature groups where feature groups were ranked by minimum redundancy - maximum relevance (MRMR) and top feature groups were selected by ridge regression, and predictive value was assessed on these top features and their associations with outcomes were examined both in NEPTUNE as training and internal validation and in CureGN as external validation.

This study leveraged previously collected digital kidney biopsies and clinical data from children and adults enrolled in the NEPTUNE^17^ and CureGN^18^ cohort studies, with a diagnosis of minimal change disease (MCD and MCD-like) or focal segmental glomerulosclerosis (FSGS). NEPTUNE and CureGN are multi-site observational cohort studies of patients with a kidney biopsy performed at the time of enrollment and within 5 years before enrollment, respectively. NEPTUNE participants that were also enrolled in CureGN (NEPTUNE transfers) were included only in the NEPTUNE dataset.

The following exclusion criteria were used for both cohorts: a) unavailability of outcome data, b) absence of PAS- WSIs; or c) failure of the PAS-stained WSIs to pass HistoQC-driven quality control^11,19,20^. CureGN participants with a biopsy performed >3 years prior to enrollment were excluded due to missing data between biopsy and study enrollment.

The UMICH cohort consists of paired PAS and immunohistochemistry (IHC) stained for pan cytokeratin WSIs from nephrectomies of kidney cancer patients, with tissue samples taken distally from the tumor mass (**Figure 1-b**). The UMICH PAS-WSIs were visually assessed by study pathologists for adequacy (i.e., good quality of tissue processing and staining). The PAS-IHC paired sections were co-registered and used as ground truth as part of the training of a DL algorithm for tubular segmentation (**METHODS 3)** and as reference tissue for comparison of pathomic feature values with the NEPTUNE/CureGN datasets.

### 2. Demographic, Clinical, and Visually Scored Pathology Data

As per NEPTUNE and CureGN study protocols, demographic data (age, sex, self-or parent-reported race and ethnicity) and WSIs were collected at study enrollment, and medication use, laboratory, and other clinical data at enrollment and each prospective 4–6-month study visit.

Clinical outcomes used in this study included (1) time from biopsy to disease progression, defined by at least 40% decline in estimated glomerular filtration rate (eGFR)^21,22^ with eGFR<90 mL/min/1.73m^2^ ^23^ or kidney failure (chronic dialysis, transplant, or two consecutive eGFRs <15 mL/min/1.73m^2^); and (2) time from biopsy to first complete proteinuria remission, defined by urine protein creatinine ratio (UPCR) <0.3 g/g. Due to eGFR and UPCR data not being collected comprehensively between biopsy and 6 months before study enrollment in CureGN, CureGN participants were considered to not have disease progression outcome data if enrolled 3-5 years after the kidney biopsy, or had fewer than 5 eGFR measurements available and less than 1.5 years of follow-up after study enrollment if biopsy was 6 months to 2 years before study enrollment. For complete proteinuria remission outcome, only participants whose biopsy was within 6 months before enrollment or at enrollment (i.e., incident patients) were included.

Tubulointerstitial descriptor scoring data (percentage of interstitial fibrosis, tubular atrophy, acute tubular injury, and inflammation) were extracted from the NEPTUNE database for NEPTUNE participants^24^, and scored by study pathologists for CureGN participants to harmonize visual scoring data across datasets.

### 3. Ground Truth and algorithmic segmentations

A mixture of manual, DL, and traditional image-processing-based segmentations were employed to segment the different kidney compartments, cortex, non-IFTA, pre-IFTA, and mature IFTA (definitions in **Supplementary Materials S2**), and tubular substructures (TL, TE, TN, and TBM). Each segmentation (ground truth and DL-generated) went through rigorous quality control by study pathologists (**Supplementary Material S2**). The segmentation approaches are detailed comprehensively in **Supplementary Materials S2**.

### 4. Quantitative feature extraction

From the segmented tubular sub-structures, a total of 99 tubule-level and 5 biopsy-level morphological pathomic features were extracted (**Supplementary Table 1**) and standardized based on micro-per-pixel (MPP) scaling to ensure consistent units for area (μm²) and length (μm) related features. **Supplementary Figure 2** shows the distribution of MPP values for both NEPTUNE and CureGN cohorts, slides for UMICH have the same MPP value (0.2527).

Tubule-level features were derived from both single and multiple sub-structures and included: i) Area (for TUBULE/TE+TL/TE/TL/TBM/TN, ii) Diameter (for TUBULE/TL/TE+TL), iii) Thickness (for TUBULE/TE+TL/TE/TL/TBM), iv) Smoothness (for TE+TL/TL/TBM), v) Nuclei location: the minimum distance from nuclei centroid/border to lumen/epithelium border, and vi) Inter-structural features: area ratio (density),and diameter, and thickness ratio between 2 sub-structures. Please see the feature extraction algorithms and detail in **Supplementary Material S3** and **Supplementary Table 1.**

Biopsy-level features included cortical TUBULE density, cortical TE+TL density, cortical TBM density, area ratio between TE+TL and TBM, and area ratio between TUBULE and TBM. Since each patient only has 1 WSI in our study, these 5 biopsy-level features are also patient-level features.

### 5. Statistical analysis

Demographics, clinical characteristics at biopsy, and visually scored tubulointerstitial morphology descriptors from NEPTUNE/CureGN study participants were described using mean and standard deviation (SD) or median and interquartile range (IQR) for continuous variables, and frequency for categorical variables. Event rates were calculated for clinical outcomes.

#### 5.1 Identification of clinically relevant tubular pathomic features in NEPTUNE

Using NEPTUNE data, 99 tubule-level pathomic features were aggregated to the patient-level using mean, standard deviation, skewness, and kurtosis, resulting in 396 patient-level features. Additionally, 5 biopsy (patient)- level pathomic features were included, resulting in 401 total patient-level tubular features used for subsequent analysis. Pairwise correlations between tubular pathomic features were assessed using Pearson’s correlation coefficient. Hierarchical clustering using Pearson’s correlation as the dissimilarity measure was employed to group highly correlated features together. The number of feature groups was chosen based on dendrogram height, such that for all feature groups, when conducting principal component analysis within each feature group, the first principal component (PC) explained >90% variability. Using the first PC to represent each feature group, the Minimum Redundancy Maximum Relevance (MRMR) selection method was used to rank feature groups, separately for each outcome. Finally, ridge-penalized Cox regression was used to assess predictive value when varying the number of top ranked feature groups included in the model, and the number of top feature groups predictive of each outcome was chosen based on the smallest number where the predictive value started to level off. Predictive value was measured by integrated area under the time-varying receiver operating characteristic curve (iAUC)^25^.

For each of the top selected feature groups, a representative pathomic feature was manually chosen using predetermined rules to optimize interpretability, for example, selecting mean over kurtosis for the same feature (**Supplementary Material S4**), and used for all subsequent analysis. Median and IQR were used to describe top selected features.

#### 5.2 Prognostic value of tubular pathomic features in NEPTUNE

To assess whether selected tubular pathomic features provide additional prognostic value above and beyond parameters currently used in routine clinical practice, a total of seven models were constructed for each outcome and their performances were compared. First, we built three models using conventional parameters alone, including (1) **<u>model 1</u>**: demographics (age, sex, Black race, Hispanic ethnicity) and clinical characteristics (FSGS vs MCD/MCD-Like, eGFR, and UPCR at the time of biopsy, and immunosuppressant use within 30 days before or at biopsy) only; (2) **<u>model 2:</u>** model 1 + % tubular atrophy and % acute tubular injury (visually assessed); (3) **<u>model 3</u>**: model 2 + % interstitial fibrosis and % inflammation (visually assessed);. Then, we built a model including selected tubular pathomic features only. Lastly, we added selected tubular pathomic features to each of the three conventional parameters models listed above. Only participants with complete data on all variables used in this analysis were included. Modeling was performed using ridge regression and predictive value was assessed by iAUC. iAUC was internally validated and bias-corrected using bootstrapping^26^.

Additionally, we estimated associations between each selected top tubular pathomic feature and the outcome using separate standard Cox proportional hazard regression. For each feature, three associations were estimated, including (i) unadjusted, (ii) adjusted for demographics and clinical characteristics only, and (iii) adjusted for demographics, clinical characteristics, and visually scored tubular morphology descriptors.

#### 5.3 External validation of clinically relevant tubular pathomic features in CureGN

CureGN tubule-level image feature data were similarly aggregated to the patient-level as in NEPTUNE. For external validation using CureGN data, we refit those seven ridge-penalized Cox models using the selected tubular pathomic features from NEPTUNE, again only including participants with complete data on all analysis variables, and similarly assessed predictive value using iAUC (with bias-correction using bootstrap). We also examined associations between the same selected tubular pathomic features and outcomes using CureGN data.

#### 5.4 Feature comparison between cortical subregions in NEPTUNE and reference tissue

Statistically significant features after full adjustment from the NEPTUNE cortical subregions (non-IFTA, pre-IFTA, and mature IFTA) were compared to UMICH reference tissue by analyzing the corresponding feature value distributions (**Figure 2**).

**Figure 2.**
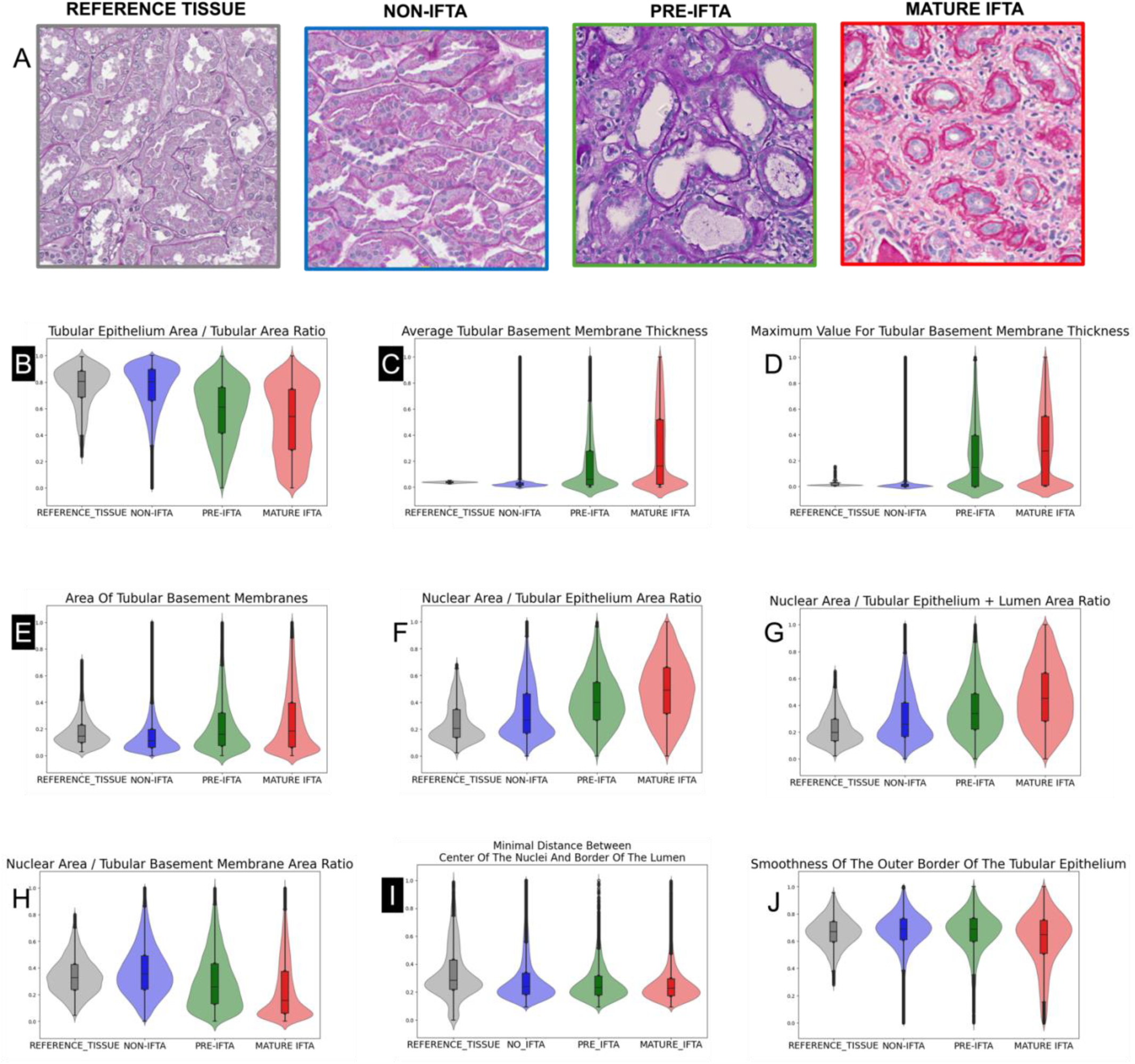
Comparison of top 9 features values between reference tissue and cortical subregions. A. Periodic Acid-Schiff (PAS)-stained Whole Slide Images (WSIs) of reference tissue from nephrectomies without interstitial fibrosis and tubular atrophy (IFTA) (grey boundary), NEPTUNE non-IFTA (blue boundary), pre-IFTA (green boundary), and mature IFTA (red boundary) regions. B-J: Violin plots illustrating the top 9 normalized feature values in reference tissue (gray violin), non-IFTA (blue violin), pre-IFTA (green violin), and mature-IFTA (red violin) regions. B: The ratio between the area of the tubular epithelium and the entire tubule is comparable across reference tissue and non-IFTA regions, although several tubules in non-IFTA regions have low values, likely reflecting acute tubular injury (simplification of the tubular epithelium). Lower values are also present in tubules in pre-IFTA (reflecting simplification of the tubular epithelium), and mature IFTA regions (reflecting simplification of the tubular epithelium in the presence of thick tubular basement membranes). C-E: The average (C) and maximum thickness (D) and area (E) of the tubular basement membranes are slightly higher and more homogeneous in reference tissue compared to non-IFTA regions, likely reflecting an overall older age for reference tissue and the presence of acute tubular injury in non-IFTA regions and increases progressively in pre-IFTA and mature IFTA. F-G: The area of tubular epithelium or tubular epithelium + lumen occupied by nuclei increases progressively from reference tissue and non-IFTA regions to pre- and mature- IFTA, indicating simplification of the tubular epithelium due to acute tubular injury, pre-atrophy, and full atrophy of tubules, respectively. H: The proportion between tubular basement membrane area and nuclear area per tubule are comparable between reference tissue and non-IFTA regions. As tubules become progressively pre- and fully atrophic the tubular basement membranes become thicker and the ratio between nuclear and tubular basement membranes’ area lower. I: Overall, the distance between the center of the tubular nuclei and the border of the lumen is greater in reference tissue compared to the 3 cortical subregions. The comparable values between non-IFTA, pre-IFTA, and mature IFTA regions reflects simplification of the tubular epithelium due to acute tubular injury, pre- and fully atrophic tubules, respectively. J: In mature IFTA, the inner boundary of the tubular basement membrane is more irregular compared to other cortical subregions and reference tissue. The 4 features (B, C, E and I) are the features which are still significant after adjustment for NEPTUNE.

## RESULTS

### 1. Study cohorts and sample collection

N=254 NEPTUNE participants (N=254 PAS-WSIs) (N=119 MCD/MCD-Like and N=135 FSGS) and N=266 CureGN participants (N=266 PAS-WSIs) (N=113 MCD/MCD-Like and N=153 FSGS) were retained in this study (**Figure 1.a**). The clinical and demographic characteristics are summarized in **Table 1**.

**Table 1:**
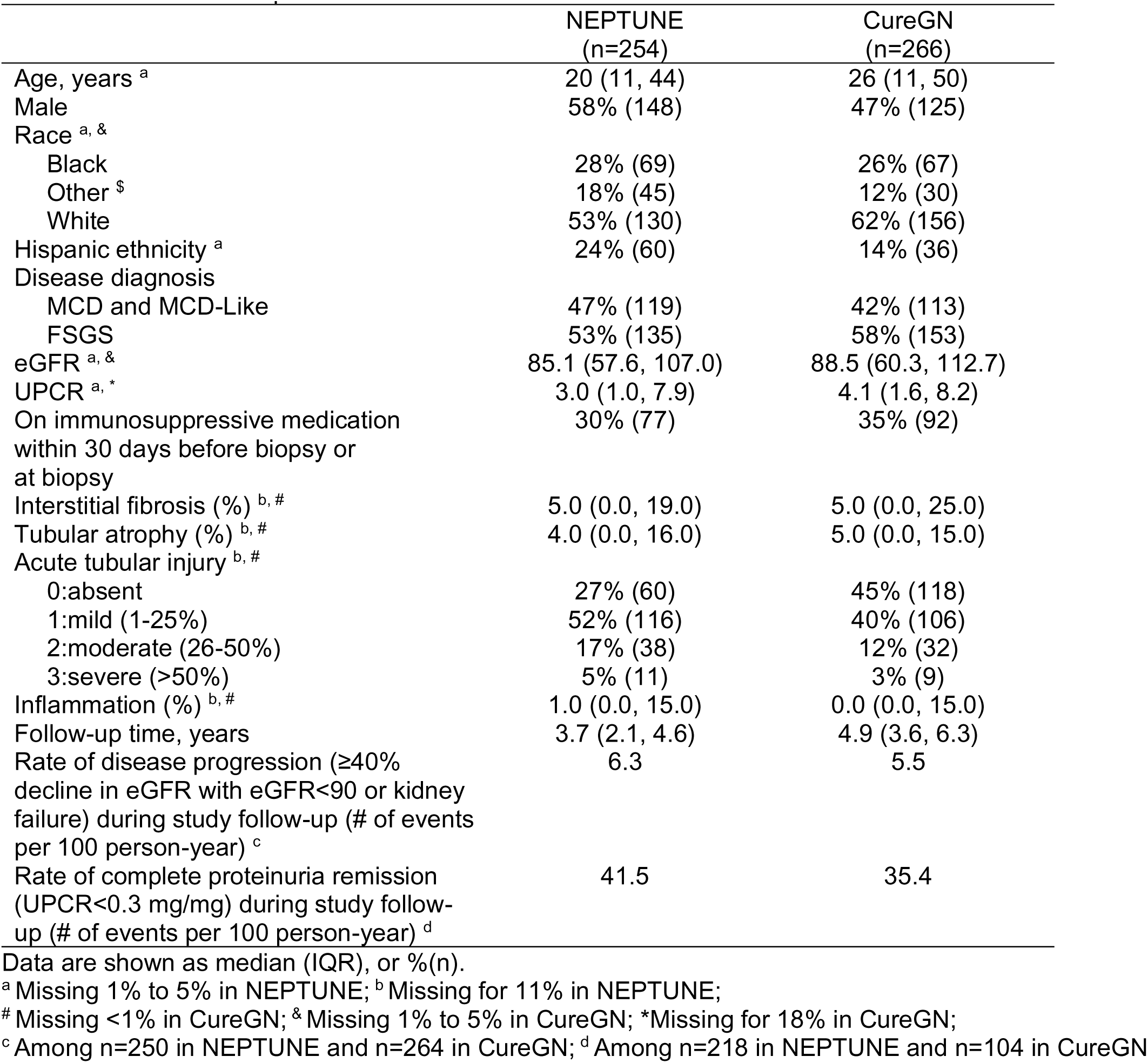
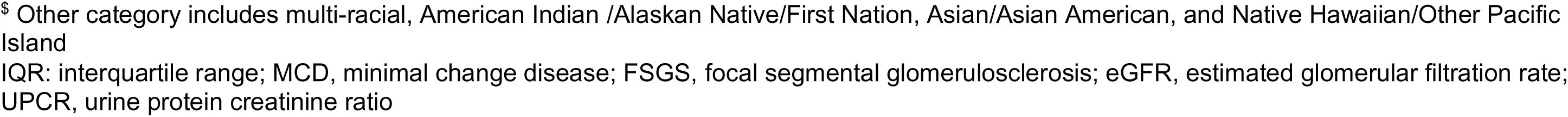
Demographics, clinical characteristics, and tubular morphologic descriptor characteristics at the time of biopsy, and study outcomes of NEPTUNE and CureGN patients.

### 2. Tubular segmentation and feature extraction

N=589,192 tubules and corresponding sub-structures (TBM, TE, TL) (N=282,658 from NEPTUNE, N=301,020 from CureGN, and N=5,514 from UMICH), and N=6,415,524 nuclei (N=3,184,105 from NEPTUNE, N= 3,179,443 from CureGN, and N=51,976 from UMICH) were segmented.

### 3. Identification of top tubular features in NEPTUNE

Among 401 patient-level tubular features, there were 779 (1%) pairwise correlations >0.90 or <-0.90. Hierarchical clustering of 401 patient-level tubular pathomic features resulted in 124 feature groups (groups vary in size from 1 to 17 features), each of which was represented by the first PC which explained >90% of its variability. Among 124 feature groups, there were only 2 (0.03%) pairwise correlations >0.90 or <-0.90 (maximum = 0.93).

Using feature groups ranked by MRMR, model performance generally increased as the number of top feature groups included in the ridge regression model increased (**Supplementary Figures 3a and 3b**). For disease progression, model performance using the top three feature groups were 98.4% of the model performance using all feature groups. For proteinuria remission, model performance using the top seven feature groups was 98.3% of the model performance using all feature groups. **Supplementary Table 2** lists these selected feature groups, including the manually chosen representative features (in **bold** text), and **Table 2** describes the distribution of the chosen representative feature from each top feature group.

**Table 2:** Tubule-level and patient-level characteristics of selected top tubular features using NEPTUNE data.

|  | Selected<br>for disease<br>progression | Selected<br>for<br>complete<br>remission | Tubule-level<br>Median (IQR) | Selected<br>patient-level<br>summary<br>statistic | Patient-level<br>Median (IQR) |
| --- | --- | --- | --- | --- | --- |
| Ratio between the area of the tubular epithelium and the area of the tubule | X |  | 0.78 (0.68, 0.85) | Mean | 0.75 (0.69, 0.79) |
| Area of the tubular basement membranes | X |  | 224.70 (155.41, 348.16) | Mean | 293.93 (252.62, 351.89) |
| Minimal distance between the center of the nuclei and the border of the tubular lumen | X | X | 2.87 (2.23, 4.04) | Mean | 3.37 (2.99, 3.88) |
| Average thickness of the tubular basement membranes |  | X | 1.06 (1.03, 1.14) | Standard Deviation | 0.39 (0.10, 1.00) |
| Ratio between the area of the nuclei and the area of the tubular epithelium |  | X | 0.16 (0.10, 0.27) | Skewness | 1.02 (0.81, 1.39) |
| Ratio between the area of the nuclei and the area of the tubular epithelium + lumen |  | X | 0.14 (0.09, 0.22) | Skewness | 1.21 (0.93, 1.63) |
| Maximum thickness of the tubular basement membranes |  | X | 1.37 (1.25, 1.50) | Skewness | 5.70 (3.00, 8.69) |
| Smoothness of the outer border of the tubular epithelium (Ramer-Douglas-Peucker epsilon = 10) |  | X | 0.94 (0.94, 0.95) | Skewness | -1.23 (-1.78, -0.65) |
| Ratio between the area of the nuclei and the area of the tubular basement membrane |  | X | 1.18 (0.77, 1.66) | Standard Deviation | 0.62 (0.55, 0.71) |

### 4. Prognostic value of selected tubular pathomic features in NEPTUNE

For the disease progression outcome, including only the top three tubular pathomic features in the model, the bias-corrected and internally validated iAUC for prognosticating outcome was 0.749 (**Table 3**). After adding these features to demographic and clinical characteristics, iAUC increased from 0.753 to 0.811. Similarly, adding these tubular pathomic features to models that included all visually scored tubulointerstitial morphology descriptors increased iAUC from 0.760 to 0.805. In standard Cox regression models, all three features had statistically significant associations with disease progression, even in fully adjusted models (**Table 4a**). Every 100 unit increase in mean TBM area was associated with 2.09 (95% CI: 1.46-3.00) times higher adjusted hazards of disease progression. Every 0.1 unit increase in mean TE: Tubule area ratio and every 1 unit increase in mean nuclei to lumen centroid distance minimum was associated with 66% (95% CI: 39%-81%) and 68% (95% CI: 37%-83%) lower adjusted hazards of disease progression, respectively.

**Table 3:** Comparison of prediction accuracy (iAUC) of clinical outcomes between conventional measures and tubular pathomic features using NEPTUNE data.

| Variables included in models |  |  | Outcomes |  |  |  |
| --- | --- | --- | --- | --- | --- | --- |
| Demographics +<br>Clinical<br>characteristics | % Tubular atrophy +<br>% Acute tubular<br>injury | % Interstitial fibrosis +<br>% Inflammation | Disease Progression<br>(n=215 participants) |  | Proteinuria Remission<br>(n=189 participants) |  |
|  |  |  | **Without**<br>MRMR top 3<br>tubular<br>features | **With**<br>MRMR top 3<br>tubular<br>features | **Without**<br>MRMR top 7<br>tubular<br>features | **With**<br>MRMR top 7<br>tubular<br>features |
|  |  |  |  | 0.749 |  | 0.724 |
| X |  |  | 0.753 | 0.811 | 0.738 | 0.772 |
| X | X |  | 0.767 | 0.810 | 0.742 | 0.760 |
| X | X | X | 0.760 | 0.805 | 0.750 | 0.762 |

**Table 4:** Associations between top tubular features and clinical outcomes in NEPTUNE from Cox proportional hazards models. Note that all top features reflected tubule-level features that were aggregated to the patient-level using summary statistics. Therefore the “mean,” “standard deviation,” or “skewness” prefixes in the feature name refers to the mean, standard deviation, or skewness across all tubules within a patient, respectively, whereas features with “minimum”, “maximum” or “average” in the middle of the feature name refers to the minimum or average of multiple measurements taken within a tubule, respectively. Demographics and clinical characteristics included age, sex, black race, Hispanic ethnicity, FSGS vs. MCD, eGFR at biopsy, UPCR at biopsy, and immunosuppressant use at biopsy. Pathology scoring variables included tubular atrophy, acute tubular injury, mononuclear WBC, and interstitial fibrosis.

|  | Unadjusted |  |  | Adjusted for demographics and clinical characteristics |  |  | Adjusted for demographics, clinical characteristics, and pathology scoring variables* |  |  |
| --- | --- | --- | --- | --- | --- | --- | --- | --- | --- |
|  | HR | 95% CI | p-value | HR | 95% CI | p-value | HR | 95% CI | p-value |
| Mean of ratios between the area of the tubular epithelium and the area of the tubule (per 0.1) | 0.37 | (0.26, 0.54) | <0.0001 | 0.27 | (0.16, 0.46) | <0.0001 | 0.34 | (0.19, 0.61) | 0.0003 |
| Mean of area of tubular basement membrane (per 100) | 1.88 | (1.51, 2.34) | <0.0001 | 1.88 | (1.41, 2.51) | <0.0001 | 2.09 | (1.46, 3.00) | 0.0001 |
| Mean of minimal distance between the center of the nuclei and the border of the tubular lumen (per 1) | 0.33 | (0.19, 0.57) | 0.0001 | 0.29 | (0.16, 0.55) | 0.0001 | 0.32 | (0.17, 0.63) | 0.0009 |

b) Complete remission
|  | Unadjusted |  |  | Adjusted for demographics and clinical characteristics |  |  | Adjusted for demographics, clinical characteristics, and pathology scoring variables* |  |  |
| --- | --- | --- | --- | --- | --- | --- | --- | --- | --- |
|  | HR | 95% CI | p-value | HR | 95% CI | p-value | HR | 95% CI | p-value |
| Standard deviation of average thickness of the tubular basement membranes (per 0.1) | 0.90 | (0.87, 0.94) | <0.0001 | 0.91 | (0.87, 0.96) | 0.0011 | 0.93 | (0.87, 1.01) | 0.0706 |
| Skewness of ratios between the area of the nuclei and the area of the tubular epithelium (per 10) | 0.26 | (0.05, 1.36) | 0.1118 | 0.70 | (0.15, 3.30) | 0.6482 | 0.76 | (0.20, 2.85) | 0.6822 |
| Skewness of ratios between the area of the nuclei and the area of the tubular epithelium + lumen (per 1) | 0.88 | (0.75, 1.03) | 0.1141 | 0.98 | (0.86, 1.13) | 0.8106 | 0.98 | (0.87, 1.10) | 0.7067 |
| Skewness of maximum thickness of the tubular basement membranes (per 1) | 1.09 | (1.05, 1.13) | <0.0001 | 1.04 | (0.98, 1.09) | 0.1930 | 1.02 | (0.96, 1.08) | 0.5773 |
| Skewness of smoothness of the outer border of the tubular epithelium (Ramer-Douglas-Peucker epsilon = 10) (per 1) | 1.39 | (1.14, 1.71) | 0.0013 | 1.16 | (0.94, 1.43) | 0.1666 | 1.14 | (0.92, 1.42) | 0.2219 |
| Standard deviation of ratio between the area of the nuclei and the area of the tubular basement membrane (per 0.1) | 0.78 | (0.68, 0.90) | 0.0006 | 0.89 | (0.77, 1.02) | 0.0970 | 0.99 | (0.84, 1.16) | 0.9037 |
| Mean of minimal distance between the center of the nuclei and the border of the tubular lumen (per 1) | 1.69 | (1.31, 2.19) | 0.0001 | 1.46 | (1.10, 1.94) | 0.0081 | 1.43 | (1.05, 1.94) | 0.0228 |

For the complete proteinuria remission outcome, including only the top seven tubular pathomic features in the model, the bias-corrected and internally validated iAUC for prognosticating outcome was 0.724 (**Table 3**). Adding these features to demographic and clinical characteristics increased iAUC from 0.738 to 0.772. Adding these tubular pathomic features to models that included visually scored tubulointerstitial morphology descriptors increased iAUC from 0.750 to 0.762. After adjustment for demographics, clinical characteristics, and visually scored tubulointerstitial morphology descriptors, only one tubular pathomic feature had a statistically significant association with complete remission (**Table 4b**). Every 1 unit increase in mean nuclei to lumen centroid distance minimum was associated with 1.43 (95% CI: 1.05-1.94) times higher adjusted hazard of complete remission.

### 5. External validation of top tubular pathomic features in CureGN

In CureGN, including only the top three and top seven tubular pathomic features had an iAUC of 0.674 and 0.763 for disease progression and complete proteinuria remission outcomes, respectively (**Table 5**), as compared with 0.749 and 0.724 in NEPTUNE (**Table 3**). Adding top tubular pathomic features to the models that included demographics and clinical characteristics increased iAUC from 0.761 to 0.769 for disease progression, and increased iAUC from 0.779 to 0.788 for complete proteinuria remission. Adding these features to the model that included visually scored tubulointerstitial morphology descriptors did not increase iAUC for disease progression and increased iAUC from 0.778 to 0.787 for complete proteinuria remission. In standard Cox regression models, two of the three top features had statistically significant associations with disease progression in models adjusting for only demographics and clinical characteristics (**Table 6a**). However, when additionally adjusting for all visually scored tubulointerstitial morphology descriptors, none remained significant, although the hazard ratios were relatively large (HR [95% CI] = 0.74 [0.38, 1.47] for mean TE: Tubule area ratio, and 1.43 [0.94, 2.18] for mean TBM Area). For complete proteinuria remission, every 0.1 unit increase in the standard deviation of TBM thickness average was associated with 14% lower adjusted hazard of complete remission (95% CI: 0%-26%) (**Table 6b**).

**Table 5:** Comparison of prediction accuracy (iAUC) of clinical outcomes between conventional measures and tubular pathomic features using CureGN data.

| Variables included in models |  |  | Outcomes |  |  |  |
| --- | --- | --- | --- | --- | --- | --- |
| Demographics +<br>Clinical<br>characteristics | % Tubular atrophy +<br>% Acute tubular<br>injury | % Interstitial fibrosis +<br>% Inflammation | Disease Progression<br>(n=197 participants) |  | Proteinuria Remission<br>(n=87 participants) |  |
|  |  |  | **Without**<br>MRMR top 3<br>tubular<br>features | **With**<br>MRMR top 3<br>tubular<br>features | **Without**<br>MRMR top 7<br>tubular<br>features | **With**<br>MRMR top 7<br>tubular<br>features |
|  |  |  |  | 0.674 |  | 0.763 |
| X |  |  | 0.761 | 0.769 | 0.779 | 0.788 |
| X | X |  | 0.793 | 0.786 | 0.790 | 0.792 |
| X | X | X | 0.786 | 0.780 | 0.778 | 0.787 |

**Table 6:** Associations between top tubular features and clinical outcomes in CureGN from Cox proportional hazards models. Note that all top features reflected tubule-level features that were aggregated to the patient-level using summary statistics. Therefore the “mean,” “standard deviation,” or “skewness” prefixes in the feature name refers to the mean, standard deviation, or skewness across all tubules within a patient, respectively, whereas features with “minimum”, “maximum” or “average” in the middle of the feature name refers to the minimum or average of multiple measurements taken within a tubule, respectively. Demographics and clinical characteristics included age, sex, black race, Hispanic ethnicity, FSGS vs. MCD, eGFR at biopsy, UPCR at biopsy, and immunosuppressant use at biopsy. Pathology scoring variables included tubular atrophy, acute tubular injury, mononuclear WBC, and interstitial fibrosis.

|  | Unadjusted |  |  | Adjusted for demographics and clinical characteristics |  |  | Adjusted for demographics, clinical characteristics, and pathology scoring variables* |  |  |
| --- | --- | --- | --- | --- | --- | --- | --- | --- | --- |
|  | HR | 95% CI | p-value | HR | 95% CI | p-value | HR | 95% CI | p-value |
| Mean of ratios between the area of the tubular epithelium and the area of the tubule (per 0.1) | 0.32 | (0.23, 0.46) | <0.0001 | 0.54 | (0.33, 0.88) | 0.0141 | 0.74 | (0.38, 1.47) | 0.3925 |
| Mean of area of the tubular basement membranes (per 100) | 2.47 | (1.89, 3.21) | <0.0001 | 1.84 | (1.28, 2.64) | 0.0010 | 1.43 | (0.94, 2.18) | 0.0928 |
| Mean of minimal distance between the center of the nuclei and the border of the tubular lumen (per 1) | 0.90 | (0.64, 1.28) | 0.5591 | 1.04 | (0.70, 1.56) | 0.8487 | 1.11 | (0.68, 1.79) | 0.6797 |

b) Complete remission
|  | Unadjusted |  |  | Adjusted for demographics and clinical characteristics |  |  | Adjusted for demographics, clinical characteristics, and pathology scoring variables* |  |  |
| --- | --- | --- | --- | --- | --- | --- | --- | --- | --- |
|  | HR | 95% CI | p-value | HR | 95% CI | p-value | HR | 95% CI | p-value |
| Standard deviation of average thickness of the tubular basement membranes (per 0.1) | 0.85 | (0.79, 0.92) | <0.0001 | 0.84 | (0.74, 0.95) | 0.0045 | 0.86 | (0.74, 1.00) | 0.0435 |
| Skewness of ratios between the area of the nuclei and the area of the tubular epithelium (per 10) | 0.41 | (0.14, 1.24) | 0.1163 | 0.73 | (0.27, 2.03) | 0.5525 | 0.58 | (0.20, 1.63) | 0.3005 |
| Skewness of ratios between the area of the nuclei and the area of the tubular epithelium + lumen (per 1) | 0.93 | (0.84, 1.02) | 0.1406 | 0.97 | (0.89, 1.06) | 0.5643 | 0.96 | (0.88, 1.05) | 0.3586 |
| Skewness of maximum thickness of the tubular basement membranes (per 10) | 1.69 | (1.08, 2.63) | 0.0205 | 0.55 | (0.25, 1.21) | 0.1401 | 0.54 | (0.23, 1.25) | 0.1490 |
| Skewness of smoothness of the outer border of the tubular epithelium (Ramer-Douglas-Peucker epsilon = 10) (per 1) | 1.42 | (1.15, 1.76) | 0.0010 | 1.01 | (0.71, 1.45) | 0.9361 | 0.91 | (0.64, 1.30) | 0.6082 |
| Standard deviation of ratio between the area of the nuclei and the area of the tubular basement membrane (per 0.1) | 0.77 | (0.59, 1.00) | 0.0463 | 1.00 | (0.73, 1.37) | 0.9867 | 1.19 | (0.84, 1.68) | 0.3344 |
| Mean of minimal distance between the center of the nuclei and the border of the tubular lumen (per 1) | 1.23 | (0.90, 1.69) | 0.1904 | 0.69 | (0.41, 1.16) | 0.1583 | 0.67 | (0.39, 1.16) | 0.1503 |

### 6. Distributions of top tubular pathomic features within cortical sub-regions and comparison to reference tissue

Overall, the tubular features values in non-IFTA regions in NEPTUNE MCD/FSGS were comparable to non-IFTA cortical regions from nephrectomies, except for nuclei: TE area ratio, nuclei: TE+TL area ratio, and the distance between the center of the nuclei and the border of the TL (luminal border of the TE) (**Figure 2 F, G, and I)**. Minimal difference was also noted in overall TBM thickness (**Figure 2>C**), likely reflecting variations across different age intervals. Specifically, individuals younger than 18 years old displayed lower feature value compared to those older than 18, suggesting potential feature variations between children and adults (**Figure 4**). When tubular feature values were compared across NEPTUNE cortical subregions, TBM thickness and area values progressively increase from non-IFTA, to pre- and mature IFTA regions (**Figure 2 C-E>).** As tubules progress toward mature atrophy, the ratio between nuclei and TE, TE+TL increases, while the nuclear area ratio to TBM decreases, which are due to TE simplification and TBM thickening (**Figure 2 F-H and Supplementary Figure 4)**. Notably, the distance between the center of the nuclei and the border of the TL (luminal border of the TE) is comparable across NEPTUNE FSGS/MCD cortical regions probably due to acute tubular injury present in non-IFTA regions. (**Figure 3 – Tubular pathway to atrophy**). These features can also provide meaningful differentiation between patients’ outcomes with more interpretability (**Figure 5**).

**Figure 3:**
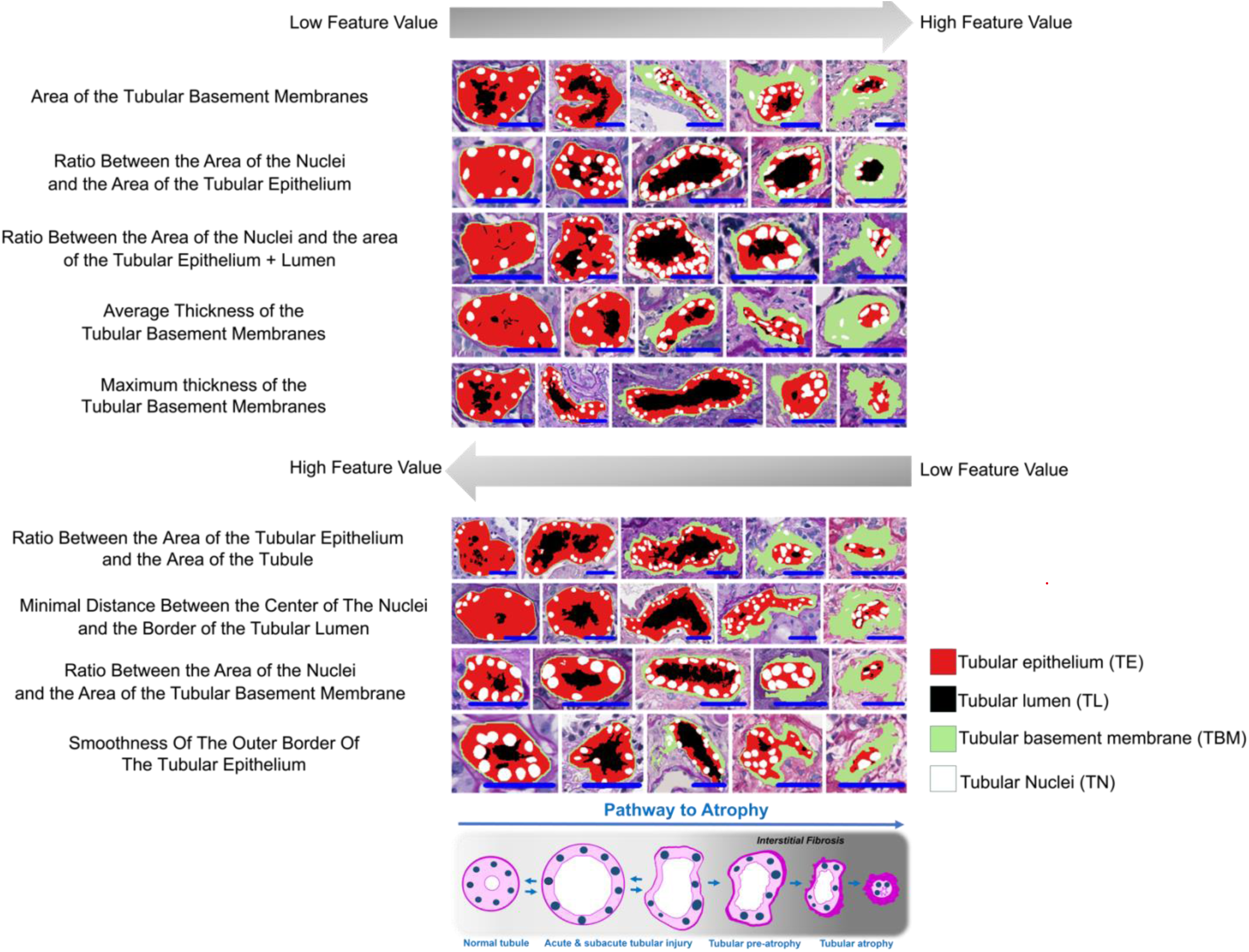
Illustrations of tubular phenotypes for the top 9 predictive features with the segmentation results: tubular epithelium in red, tubular lumen in black, tubular basement membrane in green, and tubular nuclei in white. **(top panel)** The initial five features are displayed in a sequence from left to right, illustrating a progression from lower to higher values of the feature. **(middle panel)** For the next four features, the sequence is arranged from left to right, depicting a gradient from higher to lower expression of the **(lower panel)** Cartoon illustrating the spectrum the change from normal to atrophy. A blue scale bar, located in the bottom right corner of each tubule image, represents a length of 40 micrometers.

**Figure 4.**
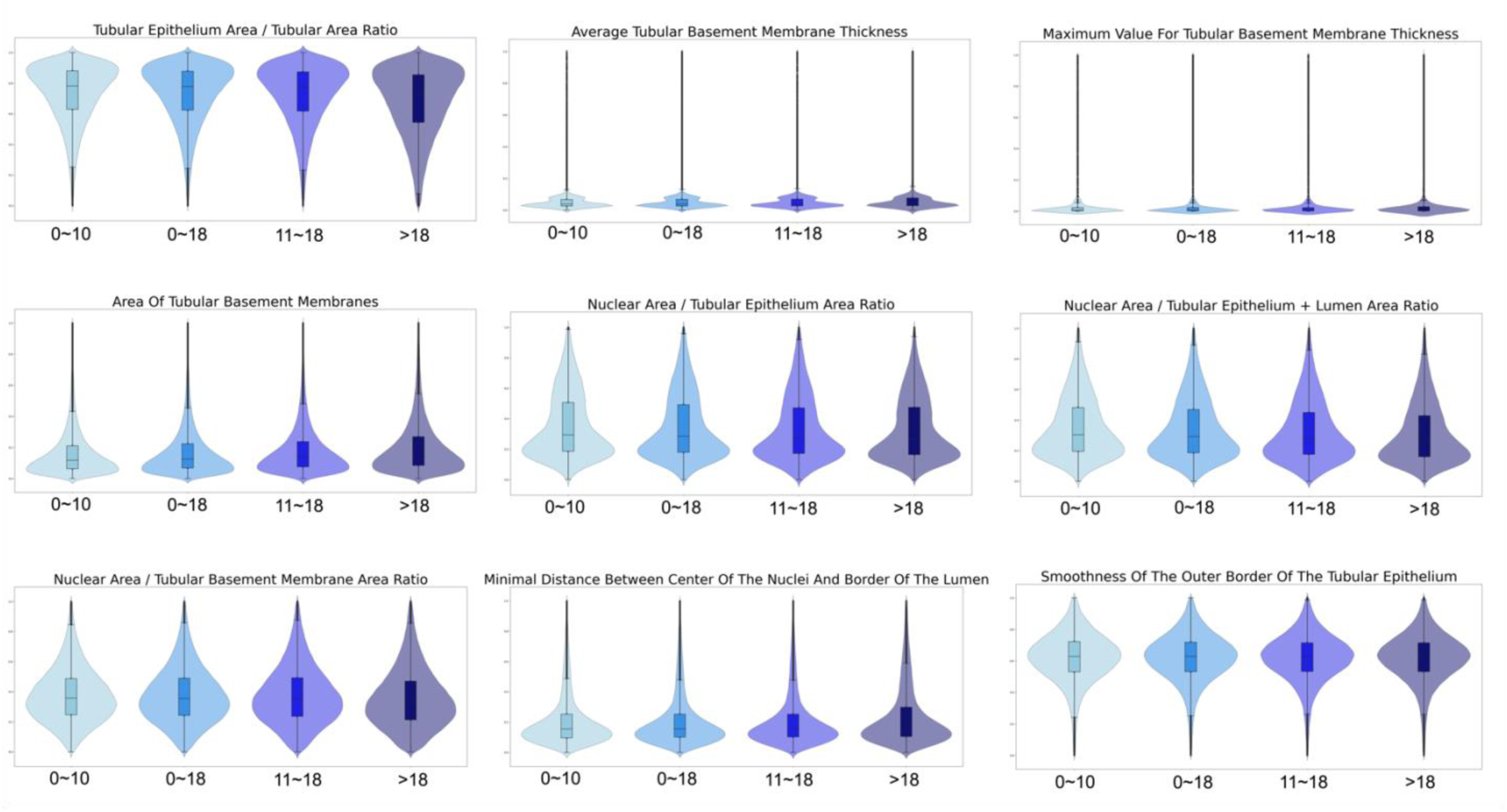
Feature comparison for NEPTUNE at different age intervals. For each violin plot, from left to right, it represents feature values in non-IFTA regions for patients with age 0∼10, 0∼18, 11∼18 and greater than 18 respectively.

**Figure 5.**
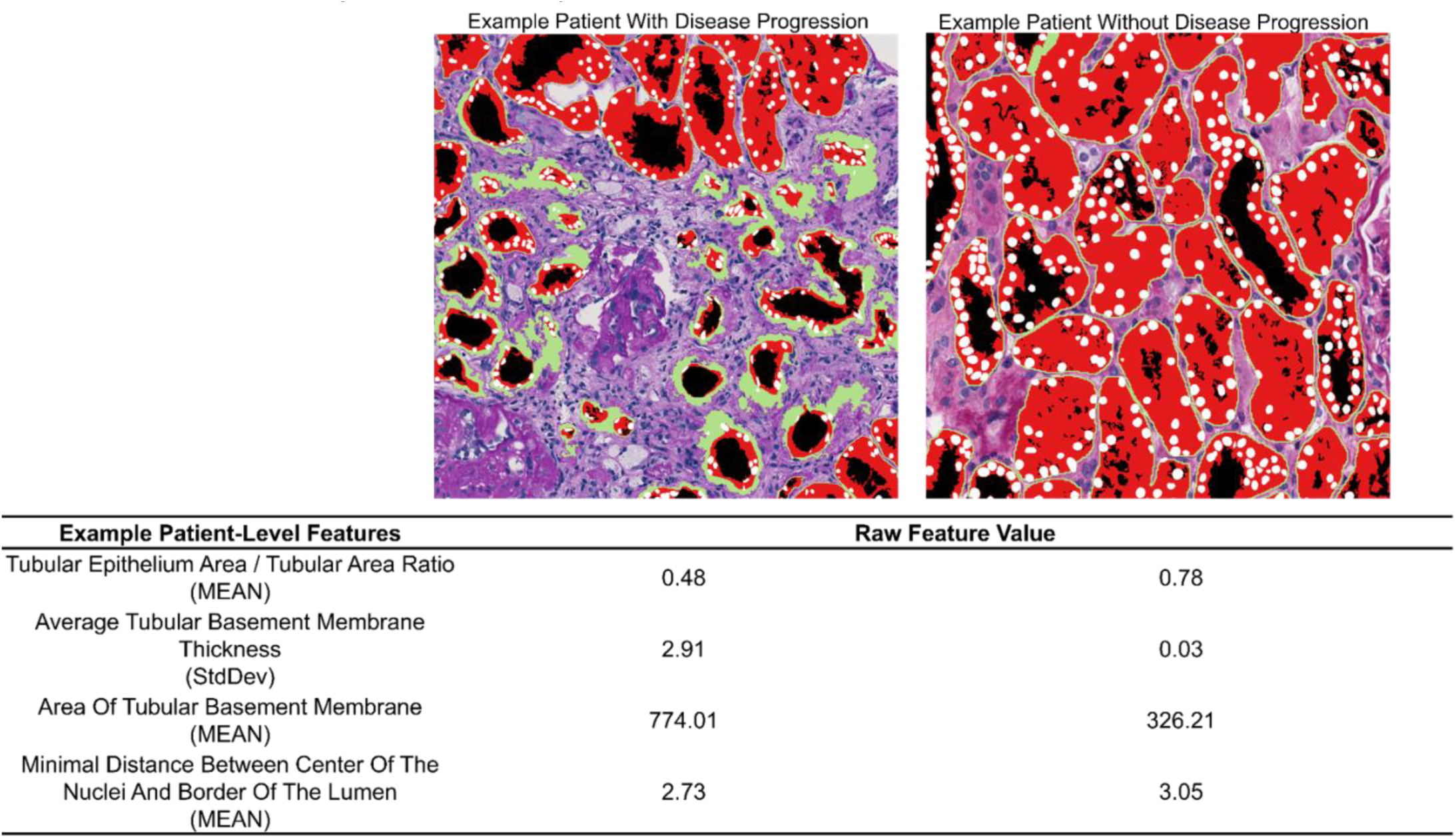
NEPTUNE patient-level feature comparison. Left patient is one patient with disease progression while the right patient is the one without disease progression. The table below shows the raw feature values for the 4 features which are still significant after adjustment between these 2 patients.

## Discussion and Conclusion

In this study, we have developed a suite of computational image analysis and machine learning algorithms aimed at identifying interpretable image-based biomarkers for renal tubules and studied the spatial relationship of tubules with interstitial scarring. We have shown that these biomarkers are prognostic of disease progression and complete remission in the NEPTUNE MCD and FSGS participants above and beyond conventional approaches. The study further delves into the evolution of tubular changes from normal to kidney scarring, by comparing tubular features between reference tissue and NEPTUNE WSIs, as well as, across different cortical subregions (non-IFTA, pre-IFTA, and mature IFTA regions).

In contrast to conventional visual scoring that is based on a semiquantitative metric (mild, moderate, and severe) to assess the presence of interstitial scarring (IFTA)^27^, and carries a variable intra- and inter-reader ^8,28^, our inherently more precise and reproducible computational approach captures the entire spectrum of tubular change and their nuanced spatial relationship with normal tissue and emerging interstitial fibrosis. For example, although TBM thickness is traditionally regarded as the visual biomarker for tubular atrophy, we were able to measure various additional TBM characteristics, and other morphological tubular features, that not only indicate atrophy but also demonstrated prognostic ability for disease progression (**Figure 3**). Similarly, we were able to measure characteristics of the tubules that reflect acute/subacute tubular injury, such as distance from the nuclear centroid to the border of the lumen and the area ratio between TE and TUBULE.

To deepen our understanding of tubular changes across normal and scarred parenchyma, we compared the top prognostic features between the reference tissues and the NEPTUNE cohort across 3 cortical subregions and demonstrated that they reveal the continuum of tubular morphological changes across non-, pre-, and mature IFTA regions. For example, we can trace an atrophy progression pathway from reference tissue through non-IFTA, pre-IFTA and mature IFTA, and the progression of tubular changes from normal to acute/subacute, and then to pre-atrophic and finally atrophic tubules. For example, in reference tissue and non-IFTA regions, the area ratio between TE and TUBULE has the higher values, with a decreasing trend progressing toward pre-IFTA and then to mature IFTA. The higher feature value in reference tissue and non-IFTA regions indicates the presence of non-simplified TE and thin TBMs (reflected by low values for area and average and maximum thickness of TBMs), consistent with normal or near to normal morphological pattern. Conversely, in pre-IFTA regions, the lower area ratio between TE and TUBULE and the lower distance of nuclear centroid to lumen border represents a trend toward simplification of TE. Since TE simplification is generally observed in acute/subacute tubular injury, one could speculate that a similar process may also be part of the pathway to atrophy when simplification occurs in the presence of interstitial fibrosis. In mature IFTA, where tubules display significantly increasing of TBM thickness and area, the lowest area ratio between TE and TUBULE is observed (refer to **Figure 2 and 3**). Notably, nephrectomies used as reference tissue are generally selected based on normal clinical parameters but may still exhibit some degree of IFTA. To obtain a more precise baseline for tubular morphometry measurements, not only did we use nephrectomies from individuals with normal clinical profiles, but we also selected tissue portions that appeared normal or near to normal to the pathologist’s eye.

In the broader kidney biomarker space, previous studies have shown the clinical relevance of image-based pathomic features extracted from peritubular capillary^7^ and glomeruli^1^. Previous work also extracted features from tubules as a unit^1,10,32^, but they were mainly focused on more basic features such as tubular size or diameter. Our scope was much broader, encompassing the development of a suite of segmentation and quantification approaches for a greater number of renal tubular structures, including epithelium, lumen, basement membranes, nuclei, and the epithelium plus lumen profile. We also employed extensive quality control to each segmentation, which was visually reviewed and manually curated when needed, to assure the highest quality prior to feature extraction. By producing visually verifiable segmentation results and biologically interpretable features, such as the area ratio between TN and TBM, our approach has the potential to reduce the inherent issues of interpretability, uncertainty, and the unknown aspects typically associated with modern end-to-end DL biomarker^29,30^ discovery approaches, which are often deemed ‘black boxes’ due to their opaque nature. As such, we believe handcrafted approaches such as ours are more likely to have near-term clinical impact due to eased clinical adoption.

This study did have some limitations worth noting. Currently, our segmentation pipeline is not fully automatic, as segmentation algorithms, both traditional and DL, often require tuning of hyperparameters (e.g., threshold for binary processing, and area threshold for morphological postprocessing) to yield optimal results. This may have potentially introduced latent batch-effects into the extracted features associated with image characteristics (e.g., brightness, contrast variance caused during the data collection process, e.g., staining). However, this concern is mitigated by the fact that segmentation results are visually verified and corrected by study pathologists, allowing for refining results with a pathologist-in-the-loop based on their expertise. Second, there are several limitations in our statistical analysis methods. Some information may be lost when aggregating tubule-level features to patient-level features using summary statistics. We used hierarchical clustering that grouped highly correlated features together and only one representative feature was chosen in each cluster or group of features. However, even highly correlated features can sometimes have different biological mechanisms that may not be captured in our analysis. Third, our study did not demonstrate that the tubular features have the same added prognostic value of clinical outcome in CureGN participants, highlighting that, despite similarity in the diagnosis and some clinical and demographic characteristics, these two cohorts are not entirely directly comparable. Future studies aim to use different datasets and expand the CureGN dataset to better investigate this phenomenon. Future work also aims to study the integration of spatial transcriptomics to help elucidate the biological relevance of these pathomic features and the mechanisms responsible for the development of kidney scarring.

In conclusion, quantitative, reproducible, computationally derived tubular features can enhance our ability to prognosticate clinical outcome beyond traditionally used parameters and can enable a more assertive stratification of patients for prognosis prediction.

## Data Availability

All data produced in the present study are available upon reasonable request to the authors.

## Acknowledgment

Research reported in this publication was supported by

1. the National Institute of Health (NIH) under the following awards: i) by the National Institute of Diabetes and Digestive and Kidney Diseases (NIDDK) under the award number 2R01DK118431-04; ii) the National Cancer Institute (NCI) under award numbers R01LM013864, R01CA249992-01A1, R01CA202752-01A1, R01CA208236-01A1, R01CA216579-01A1, R01CA220581-01A1, R01CA257612-01A1, 1U01CA239055-01, 1U01CA248226-01, 1U54CA254566-01, and 1U01DK133090; iii) the National Heart, Lung and Blood Institute under award numbers 1R01HL15127701A1, R01HL15807101A1; iv) the National Institute of Biomedical Imaging and Bioengineering under award number 1R43EB028736-01; and v) the National Center for Research Resources under award number 1 C06 RR12463-01,
2. VA Merit Review Award IBX004121A from the United States Department of Veterans Affairs Biomedical Laboratory Research and Development Service.
3. the Office of the Assistant Secretary of Defense for Health Affairs, through i) the Breast Cancer Research Program (W81XWH-19-1-0668), ii) the Prostate Cancer Research Program (W81XWH-15-1-0558, W81XWH-20-1-0851), iii) the Lung Cancer Research Program (W81XWH-18-1-0440, W81XWH-20-1-0595), iv) the Peer Reviewed Cancer Research Program (W81XWH-18-1-0404, W81XWH-21-1-0345). 4) the Kidney Precision Medicine Project (KPMP) Glue Grant and sponsored research agreements from Bristol Myers-Squibb, Boehringer-Ingelheim, and Astrazeneca.
4. The Nephrotic Syndrome Study Network (NEPTUNE) is part of the Rare Diseases Clinical Research Network (RDCRN), which is funded by the National Institutes of Health (NIH) and led by the National Center for Advancing Translational Sciences (NCATS) through its Division of Rare Diseases Research Innovation (DRDRI). NEPTUNE is funded under grant number U54DK083912 as a collaboration between NCATS and the National Institute of Diabetes and Digestive and Kidney Diseases (NIDDK). Additional funding and/or programmatic support is provided by the University of Michigan, NephCure Kidney International, Alport Syndrome Foundation, and the Halpin Foundation. RDCRN consortia are supported by the RDCRN Data Management and Coordinating Center (DMCC), funded by NCATS and the National Institute of Neurological Disorders and Stroke (NINDS) under U2CTR002818.
5. Additional support was also provided by NephCure and the Henry E. Haller, Jr. Foundation.
6. Funding for the CureGN consortium is provided by U24DK100845 (formerly UM1DK100845), U01DK100846 (formerly UM1DK100846), U01DK100876 (formerly UM1DK100876), U01DK100866 (formerly UM1DK100866), and U01DK100867 (formerly UM1DK100867) from the NIDDK/NIH. Patient recruitment is supported by NephCure. Dates of funding for first phase of CureGN was 9/16/2013-5/31/2019.
7. This work was supported by the National Science Foundation Graduate Research Fellowship [DGE-2236662 to JR]

Members of the Nephrotic Syndrome Study Network (NEPTUNE)

NEPTUNE Collaborating Sites

*Atrium Health Levine Children’s Hospital, Charlotte, SC*: Susan Massengill*, Layla Lo^#^

*Cleveland Clinic, Cleveland, OH*: Katherine Dell*, John O’Toole*, John Sedor**, Victoria Grange^#^

*Children’s Hospital, Los Angeles, CA*: Ian Macumber*, Alyssa Parry^#^

*Children’s Mercy Hospital, Kansas City, MO*: Tarak Srivastava*, Kelsey Markus^#^

*Cohen Children’s Hospital, New Hyde Park, NY*: Christine Sethna*, Suzanne Vento^#^

*Columbia University, New York, NY:* Pietro Canetta*

*Duke University Medical Center, Durham, NC:* Opeyemi Olabisi*, Rasheed Gbadegesin**, Maurice Smith^#^

*Emory University, Atlanta, GA:* Laurence Greenbaum*, Chia-shi Wang*, Emily Yun^#^

*The Lundquist Institute, Torrance, CA:* Sharon Adler*, Janine LaPage^#^

*John H Stroger Cook County Hospital, Chicago, IL:* Amatur Amarah*

*Johns Hopkins Medicine, Baltimore, MD:* Meredith Atkinson*, Ryan Hutson^#^

*Mayo Clinic, Rochester, MN:* John Lieske, Marie Hogan, Fernando Fervenza

*Medical University of South Carolina, Charleston, SC:* David Selewski*, Cheryl Alston^#^

*Montefiore Medical Center, Bronx, NY:* Kim Reidy*, Michael Ross*, Frederick Kaskel**, Patricia Flynn^#^

*New York University Medical Center, New York, NY:* Laura Malaga-Dieguez*, Olga Zhdanova**, Laura Jane Pehrson^#^, Melanie Miranda^#^

*The Ohio State University College of Medicine, Columbus, OH*: Salem Almaani*, Laci Roberts^#^

*Stanford University, Stanford, CA:* Richard Lafayette*, Shiktij Dave^#^

*Temple University, Philadelphia, PA:* Iris Lee**

*Texas Children’s Hospital at Baylor College of Medicine, Houston, TX*: Shweta Shah*, Sadaf Batla^#^ ^#^

*University Health Network Toronto:* Heather Reich*, Michelle Hladunewich**, Paul Ling^#^, Martin Romano^#^

*University of California at San Francisco, San Francisco, CA*: Paul Brakeman*, Daniel Schrader

*University of Colorado Anschutz Medical Campus, Aurora, CO*: James Dylewski* Nathan Rogers^#^

*University of Kansas Medical Center, Kansas City, KS*: Ellen McCarthy*, Catherine Creed^#^

*University of Miami, Miami, FL:* Alessia Fornoni*, Miguel Bandes^#^

*University of Michigan, Ann Arbor, MI:* Matthias Kretzler*, Laura Mariani*, Zubin Modi*, A Williams^#^, Roxy Ni^#^

*University of Minnesota, Minneapolis, MN:* Patrick Nachman*, Michelle Rheault*, Amy Hanson^#^, Nicolas Rauwolf^#^

*University of North Carolina, Chapel Hill, NC:* Vimal Derebail*, Keisha Gibson*, Anne Froment^#^, Mary Mac McGown Collie^#^

*University of Pennsylvania, Philadelphia, PA:* Lawrence Holzman*, Kevin Meyers**, Krishna Kallem^#^, Aliya Edwards^#^

*University of Texas San Antonio, San Antonio, TX*: Samin Sharma**

*University of Texas Southwestern, Dallas, TX:* Elizabeth Roehm*, Kamalanathan Sambandam**, Elizabeth Brown**, Jamie Hellwege

*University of Washington, Seattle, WA:* Ashley Jefferson*, Sangeeta Hingorani**, Katherine Tuttle^**§^, Linda Manahan ^#^, Emily Pao^#^, Kelli Kuykendall^§^

*Wake Forest University Baptist Health, Winston-Salem, NC:* Jen Jar Lin**

*Washington University in St. Louis, St. Louis, MO*: Vikas Dharnidharka*

Data Analysis and Coordinating Center: *University of Michigan:* Matthias Kretzler*, Brenda Gillespie**, Laura Mariani**, Zubin Modi**, Eloise Salmon**, Howard Trachtman**, Tina Mainieri, Gabrielle Alter, Michael Arbit, Hailey Desmond, Sean Eddy, Damian Fermin, Wenjun Ju, Maria Larkina, Chrysta Lienczewski, Rebecca Scherr, Jonathan Troost, Amanda Williams, Yan Zhai; *Arbor Collaborative for Health:* Colleen Kincaid, Shengqian Li, Shannon Li; *Cleveland Clinic:* Crystal Gadegbeku**, *Duke University:* Laura Barisoni**; John Sedor**, *Harvard University:* Matthew G Sampson**; *Northwestern University:* Abigail Smith**; *University of Pennsylvania:* Lawrence Holzman**, Jarcy Zee**

Digital Pathology Committee: Carmen Avila-Casado *(University Health Network)*, Serena Bagnasco *(Johns Hopkins University)*, Lihong Bu *(Mayo Clinic)*, Shelley Caltharp *(Emory University)*, Clarissa Cassol *(Arkana)*, Dawit Demeke *(University of Michigan)*, Brenda Gillespie *(University of Michigan)*, Jared Hassler *(Temple University)*, Leal Herlitz *(Cleveland Clinic)*, Stephen Hewitt *(National Cancer Institute)*, Jeff Hodgin *(University of Michigan)*, Danni Holanda *(Arkana)*, Neeraja Kambham *(Stanford University)*, Kevin Lemley, Laura Mariani *(University of Michigan)*, Nidia Messias *(Washington University)*, Alexei Mikhailov *(Wake Forest)*, Vanessa Moreno *(University of North Carolina)*, Behzad Najafian *(University of Washington)*, Matthew Palmer *(University of Pennsylvania)*, Avi Rosenberg *(Johns Hopkins University)*, Virginie Royal *(University of Montreal)*, Miroslav Sekulik *(Columbia University)*, Barry Stokes *(Columbia University)*, David Thomas *(Duke University)*, Ming Wu *(University of New York)*, Michifumi Yamashita *(Cedar Sinai)*, Hong Yin *(Emory University)*, Jarcy Zee *(University of Pennsylvania)*, Yiqin Zuo *(University of Miami)*. Co-Chairs: Laura Barisoni *(Duke University)*, Cynthia Nast *(Cedar Sinai)*.

*Principal Investigator; **Co-investigator; ^#^Study Coordinator; ^§^Providence Medical Research Center, Spokane, WA Last Update: 12OCT2023

