## supplementary material for "Clinical Relevance of Computationally Derived Tubular Features: Spatial Relationships and the Development of Tubulointerstitial Scarring in MCD/FSGS"

### METHODS

*S1. Demographic, Clinical, and Visually Scored Pathology Data*

1. **Self-reporting race:**
   Reporting race and ethnicity was mandated by the U.S. National Institutes of Health, consistent with the Inclusion of Women, Minorities, and Children policy.

2. **Immunosuppressive medications**:
   Immunosuppression exposure was defined as any immunosuppression use within 30 days before biopsy or at biopsy, using medication start and stop dates, as identified by study coordinators. Immunosuppression use included any combination of corticosteroids, calcineurin inhibitors, mycophenolate mofetil, cyclophosphamide, and rituximab, among others.

3. **Laboratory Data:**
   Serum creatinine and urine protein creatinine ratio (UPCR) were measured by a central laboratory from blood and urine samples collected at each study visit. Local serum creatinine and UPCR measurements were also collected at each study visit and in between study visits, including at the time of biopsy in CureGN. Estimated glomerular filtration rate (eGFR) was estimated on the basis of serum creatinine levels using the race-free CKD-Epi equation for adults 25 years or older and the Under 25 (U25) equation for children and adults <25 years old[1]. UPCR was calculated from a 24-hour urine collection, first-morning void, or random spot urine.

*S2. Ground Truth and Algorithmic Segmentations*

Described below are the approaches used to segment a) the kidney compartments: cortex, mature interstitial fibrosis and tubular atrophy (IFTA), pre-IFTA, and non-IFTA; and b) the tubular substructures: tubular lumen (TL), tubular epithelium (TE), tubular nuclei (TN), and tubular basement membrane (TBM). Pre-IFTA was defined by the presence of interstitial fibrosis containing tubules with disrupted cellular architecture, size comparable to non-atrophic tubules, and mild thickening of the TBMs. Mature IFTA was defined by the presence of interstitial fibrosis containing small tubules with very thick TBMs[2]. Non-IFTA was defined by the absence of pre- and mature IFTA.

1. **Cortex and cortical subregions:**
   QuPath[3] was used for manual segmentation of (a) the cortex in the NEPTUNE, CureGN, and UMICH WSI datasets, and for (b) pre-, mature, and non-IFTA sub-regions in NEPTUNE. UMICH PAS-WSIs were visually assessed by study pathologists and portions of cortex that were comparable in size to the average kidney biopsy and that did not contain IFTA, or inflammation, were selected and manually segmented to serve as reference tissue.

2. **Tubular Epithelium + Tubular Lumen (TE+TL):**
   U-Net (a DL model architecture)[4] was trained in a two-phase approach including IHC bootstrapping and manual segmentation followed by pathologist-in-the-loop refinement.

   2.1. *IHC Bootstrapping*:
   26 UMICH paired PAS-IHC pan-cytokeratin/ALDOB stained WSIs were used to generate ground truth for TE+TL structures. Tissue sections were first stained with PAS and scanned at 40X. The coverslip was then removed, and sections were immersed in 10 mM sodium citrate buffer (pH 6.0, ab93678, Abcam) at 95°C for 2 hours for antigen retrieval and then were incubated at 4°C overnight with the primary antibody cocktail by mixing rabbit anti-AE1/AE3 antibody (ready to use, GA05361-2, Agilent) and mouse anti-ALDOB antibody (1:900, HPA002198, Sigma). Antibody binding was detected by using secondary antibody cocktail containing goat anti-Rabbit Ig-HRP (1:50, 4010-05, Southern Biotech) and goat anti-mouse Ig-HRP (1:100, 1010-05, Southern Biotech) and betazoid DAB chromogen kit (BDB2004, Biocare Medical). N=41 regions of interest (ROIs) containing only tubules were manually selected and extracted from the PAS WSIs, and then co-registered with IHC ROIs using a cross-correlation algorithm[5,6]. IHC-stained ROIs (at 40X) were used to create binary masks which served as ground truth for the corresponding PAS ROIs (**Supplementary Figure 1-a.**). This

process involved the following steps: (1) applying binary thresholding with a threshold value of 150 to roughly delineate the TE+TL regions, (2) a blurring step using a kernel of size 5 to smooth the segmented areas; (3) a second binary thresholding of 60 was then applied to refine the segmentation results; and (4) the pixel values of the binary image were inverted, setting the TE+TL to a white pixel value (255) and all other pixels to black (0); and (5) morphological processing steps: the morphological functions from the Skimage package used included (i) hole filling (**remove_small_objects** function from the Skimage library with an area thresholding of 2000), and (ii) object removal (**remove_small_objects** function from the Skimage library with an area thresholding of 800) followed by, and (iii) erosion (employing a kernel size of 2 across 2 iterations).

*2.2.* *Manual segmentation*:

For this study we also utilized 65 ROIs from 58 PAS-WSIs from the NEPTUNE WSI dataset that were previously manually segmented by study pathologists[7] (**Supplementary Figure 1-a.**).

*2.3.* *U-Net model training:*

U-Net[4,7], a deep learning (DL) architecture, was used for the automatic segmentation tasks. The first model was trained using 4503 TE+TL structures from the 41 UMICH ROIs and 4802 TE+TL structures from the 65 NEPTUNE ROIs. Each ROI was cropped to a dimension of 3000X3000 at 40X magnification for consistency. The architecture was implemented in PyTorch with the following configuration details: (a) depth of the U-Net: 5 blocks, number of filters in the filter layer: 4; (b) patch size: 512x512, number of training batches for each epoch: 6; (c) number of training epochs: 50, the model with the lowest validation loss (10% of the patients were selected as validation set) was used to do the testing; (d) optimization algorithm: Adam; (e) data augmentation: stain normalization[8], vertical & horizontal flips, rotation & hue, saturation, and value adjustment were used during the network training process; (f) magnification used for training the model: 10X.

*2.4.* *Pathologist-in-the-loop refinement:*

An iterative training process was implemented, incorporating pathologist validation into the training set to iteratively improve the TE+TL segmentation DL model's accuracy[9]. The cortical tissue bounding box was first cropped based on the cortex annotation and downsampled to 10X magnification. Utilizing the U-Net model at previous iteration, initial TE+TL segmentations were generated on these bounding box images. Subsequently, we employed a suite of Python libraries – OpenCV, Shapely, and JSON – to translate bounding box-level binary mask annotations into WSI-level JSON files that are compatible with QuPath. A centralized OMERO server[10] was used to store the WSI datasets and enable pathologists to remotely access the WSIs. Study pathologists were provided with a QuPath project folder containing the TE+TL annotations produced by the U-Net model linked to the WSIs contained in the OMERO server. After logging into the OMERO server with the given credentials, study pathologists evaluated and refined the annotations when needed, utilizing the QuPath's annotation tools. This quality control and refinement processes involved erasing false positives, manually annotating false negatives, and disentangling 'super tubules' (tubules that are connected). These expert modifications served as final segmentations used for downstream feature extraction and feedback to enhance the model's performance in successive iterations (training details are the same as above 'U-Net model training' section).

### 3. Tubular Basement Membrane (TBM):

Two image processing approaches at 10X magnification were used to generate the TBM segmentation based on their thicknesses. The rationale for this approach is that very thick TBMs of atrophic tubules are very different in texture and contour compared to TBMs of non-atrophic tubules, thus, the approach used for the latter would have not accurately segmented the boundaries of the thick TBMs.

*3.1.* *For TBMs with normal thickness*.

Refined TE+TL segmentation boundaries were expanded using dilation (kernel size of 5, iteration number of 2). The TBMs with normal thickness were then extracted by subtracting the raw TE+TL from the dilated TE+TL boundaries using bitwise operators NOT and AND. These algorithms were implemented using the Python package OpenCV.

*3.2. For the thickened TBM*.

A structure-preserving color normalization (Vahadane)[8] was first used to normalize the original PAS ROIs using a reference image. The normalized images were then stained and deconvoluted[11] using a hematoxylin-eosin deconvolution matrix to isolate the “eosin” channel, which sufficiently highlighted the thickened TBM. This channel was subsequently used for segmentation. The image processing workflow involved initial binary processing (using a threshold of 150 or 160, which may be adjusted based on the image), followed by blurring (kernel size of 2), grayscale conversion, and a second binary thresholding (threshold of 220). Morphological postprocessing was then applied, including the removal of false positives (using the **remove_small_objects** function from the Skimage library with an area threshold of 100) and the filling of false negatives (using the **remove_small_holes** function with an area threshold of 1000). To further reduce false positives, two previously validated DL segmentation models[7] were utilized to exclude glomeruli and vessels, for example, the glomerular basement membrane was removed using bitwise operators NOT and AND (**Supplementary Figure 1. b**). The segmented areas for normal and thickened TBMs were merged to represent the TBM thickness spectrum using the bitwise operator AND. Python libraries including OpenCV, Skimage, and NumPy were employed throughout this process.

*3.3. Pathologist-in-the-loop refinement:*

A similar refinement loop process described above for the TE+TL segmentation was used for TBM segmentation.

**4. Tubular Lumen (TL):**

In our analysis, the white areas within the tubule regions are identified as the lumen. To segment the TL, we employed a sequence of image processing techniques (at 40X) beginning with grayscale conversion followed by binary thresholding, where the threshold parameter is adjusted on a case-by-case basis through visual assessment, the median value used in the study cohorts is 190. We retained only the white areas within the TE+TL structure identified by the deep learning model and refinement, using AND operators. The post-processing steps were conducted at the tubule level to refine the segmentation results. We utilized the **remove_small_objects** function from the Skimage library, setting the threshold at 10% of the area of the largest detected lumen to remove smaller, irrelevant objects. Additionally, we filled the false negatives by employing the **remove_small_holes** function, with a threshold area of 1000, to fill in any gaps within the lumen areas.

**5. Tubular Epithelium (TE):**

Segmentations for the TE were obtained by subtracting the TL from the TE+TL segmentations using bitwise operators such as NOT and AND in the OpenCV library of Python.

**6. Tubular Nuclei (TN):**

A pre-trained HoverNet[12] was used to segment nuclei at 40X on WSI-level. HoverNet is an advanced DL architecture specifically designed for nuclear segmentation and classification within histopathology WSIs and utilizes an instance-aware segmentation strategy, which distinguishes between overlapping and touching nuclei. The **remove_small_objects** function from the Skimage library was used to remove the false positive by setting the area threshold as 50.

***7.* Tubule (TUBULE):**

The segmentation for the entire tubular structure was obtained by adding TE+TL+TBM segmentations and will be referred to as TUBULE.

*S3. Quantitative feature extraction*

Feature extraction was conducted at the tubular level, where 99 tubule-level morphological features were extracted from various substructures including TE, TL, TE+TL, TBM, and TUBULE. These features encompass area, thickness/diameter, smoothness, nuclear location, density, and the ratio of thickness to diameter. TUBULEs with a TE+TL area smaller than 50 pixels were excluded from the feature extraction pipeline. All tubular substructures were stored as binary PNG files, where the masks were highlighted as white pixels (255). Structures from the same tubule shared the same file ID to facilitate easy identification. In addition to the features at the tubule level, five overarching biopsy-level features were also extracted for each patient. TraitHorizon (https://github.com/choosehappy/TraitHorizon), an open-source quality control tool was used to verify the extracted features, identify outliers and scrutinize problematic segmentation results.

1. **Area:**
   Area features were extracted from TUBULE, TE, TL, TE+TL, and TBM. The Python libraries OpenCV and NumPy were utilized for this process. Function ‘findContours’ in OpenCV (retrieval mode is cv2.RETR_TREE and approximation method is cv2.CHAIN_APPROX_SIMPLE) was used to determine each structure’s contour, and NumPy was employed to count the number of white pixels, which represents the pixel-level area value.

2. **Thickness/Diameter:**
   Thickness features were extracted from TUBULE, TE, TL, and TE+TL using Python libraries Skimage, OpenCV, and NumPy. Initially, OpenCV generated a distance transformation map[13] of the structure using the L2 norm for distance calculation and a mask size of 3. This distance transformation produced a heatmap where the pixel density represents the distance between the white pixel and the nearest zero pixel. A skeletonization method[14] was then applied to reduce the binary objects to a 1-pixel-wide representation of the structure's topology. By multiplying the distance transformation heatmap by the skeleton, the resulting pixel density along the skeleton indicated the distance from the skeleton to the structural border. Doubling this value yielded the thickness of the structure. First-order statistics (minimum, maximum, average, and standard deviation) were derived from these measurements. For further details, please see the illustration in (**Supplementary Figure 5-a**). The maximum value of the thickness was used to represent the diameter of the tubular structures representing the diameter of the largest inscribed circle of the structure (**Supplementary Figure 5-b**).

3. **Smoothness:**
   Smoothness is assessed by examining the irregularity of a structure's outer boundary. Initially, OpenCV retrieved the raw contour coordinates of the structure using the function ‘findContours’. Using the Ramer-Douglas-Peucker (RDP) algorithm[15], the contour was simplified at varying epsilon values (10, 15, 20, and 30) to reduce the number of points while preserving the overall shape. Epsilon determines the degree of simplification; a lower epsilon implies the contour remains closer to the original. Smoothness was quantified by the ratio between the arc length of the simplified contour to that of the raw contour, with a value of 1 indicating perfect correspondence (**Supplementary Figure 5-c**).

4. **Nuclear Location:**
   The minimum distance from the center or border of the TN to the border between the TE and TL was determined to assess the location of nuclei within the TE. Initially, OpenCV was utilized to capture the contour of each nucleus, for the center of TN, the center was calculated using the contour's moments[16]. For efficient determination of the nearest neighbor distances, a KD-tree was constructed with the coordinates of the nuclear center of boundaries. This tree was then queried to identify the closest point on a nucleus and calculate the distance to the border between the TE and TL (**Supplementary Figure 5-d**). The entire process leveraged Python’s OpenCV, Scipy, and Numpy libraries.

5. **Inter-structural features:**
   Area (density), diameter, and thickness (average thickness) ratio were extracted between 2 sub-structures (**Supplementary Table 1).**

6. **Biopsy-level features:**
   5 biopsy-level features including cortical TUBULE/TBM/TE+TL density. These density metrics were derived by summing the areas of the TUBULE, TBM, and TE+TL structures and then computing the ratio of these cumulative areas to the area of the cortex. The remaining two features involved the area ratios between cortical TE+TL to TBM and cortical TUBULE to TBM. These were determined by first calculating the total area of cortical TE+TL and TUBULE, respectively, and then calculating the proportional area ratios to the cortical TBM area.

*S4. Choosing a representative pathomic feature in each feature group*

Predetermined rules were used in selecting a representative pathomic feature within each of the selected top feature group: (i) if multiple tubule features were in a feature group, we selected patient level over tubule level feature, non-summarized over summarized feature, non-ratio over ratio feature, area over diameter over thickness over smoothness feature (for TUBULE, TE+TL, TL features), and centroid over border (for TN features); (ii) if multiple patient-level summary statistics for the same tubule feature, we prioritized them based on the order listed here: mean (most prioritized), max, min, standard deviation, skewness, kurtosis (least prioritized); (iii) if different tubule components for non-ratio tubule features, we selected single component over multiple components, for example, selecting TE or TL over TE + TL features, or selected larger component over smaller component, for example, selecting TUBULE over TE or TL or TBM; (iv) if different tubule components for ratio tubule features, we selected nested ratio over non-nested ratio.

**Reference – Supplementary material**

**Supplementary Figure 1** Detailed visualization of ground-truth for renal tubular structure segmentation. **a.** Creation of ground truth data for the preliminary model targeting segmentation of tubular epithelium (TE) and lumen (TL), utilizing registered immunohistochemistry (IHC) images to segment the combined tubular epithelium and lumen in the corresponding Periodic Acid-Schiff (PAS) stained images. **b.** Details of the algorithm used for segmenting the tubular basement membrane.

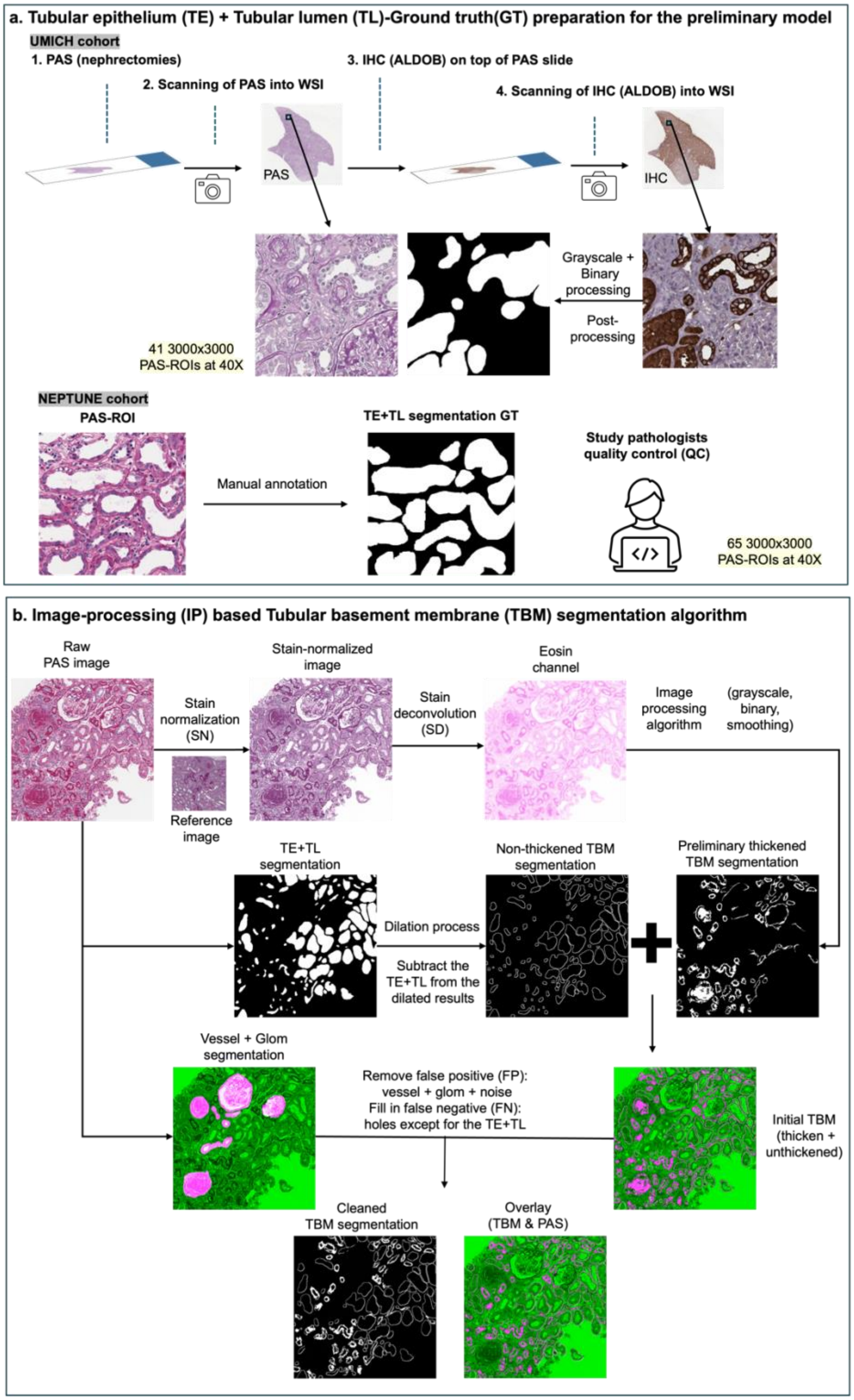

**Supplementary Figure 2** Distribution of micrometer per pixel (mpp) values for NEPTUNE and CureGN Cohorts.

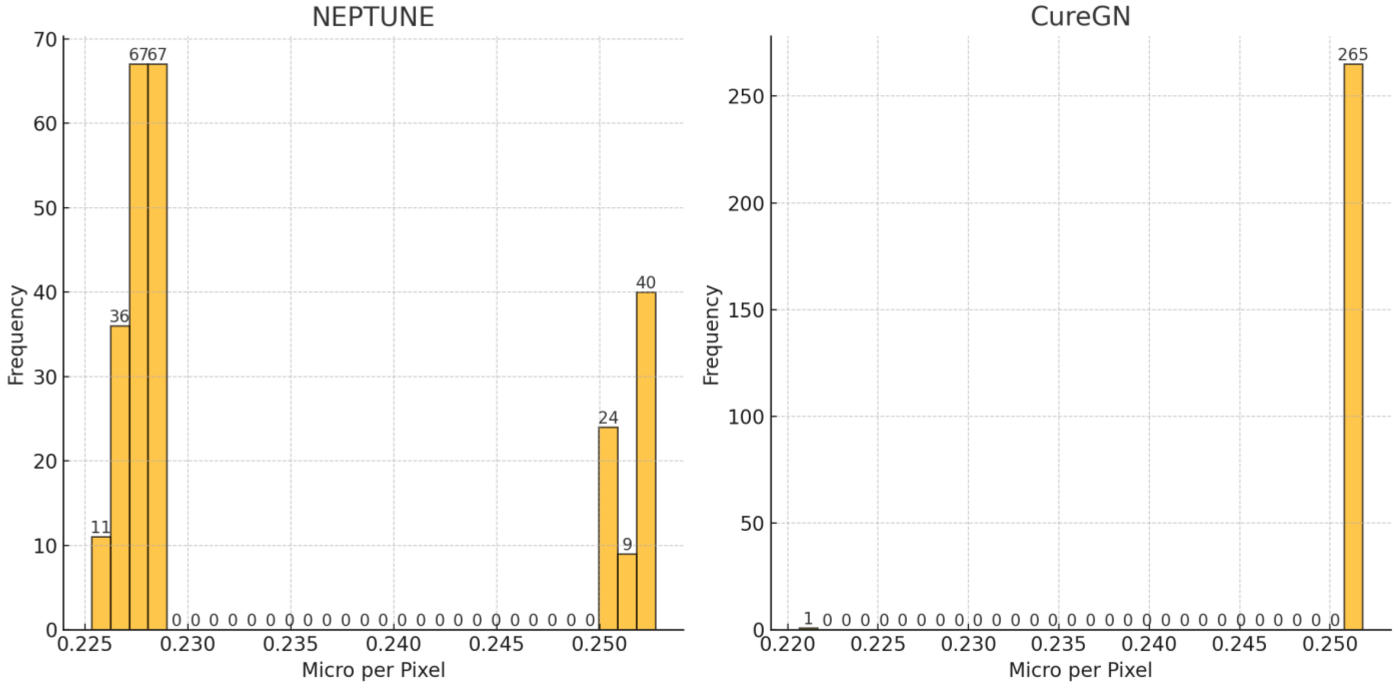

**Supplementary Figure 3:** Prediction accuracy (iAUC) from ridge regression model by varying number of top tubular feature groups included, using NEPTUNE data. Tubular feature groups were ranked by MRMR. For disease progression (figure a), including only the top three feature groups resulted in an iAUC of 0.775, which was 98.4% of 0.788, the iAUC when including all feature groups. For proteinuria remission (figure b), including the top seven feature groups resulted in an iAUC of 0.745, which was 98.3% of 0.758, the iAUC when including all feature groups.

(a) Disease progression

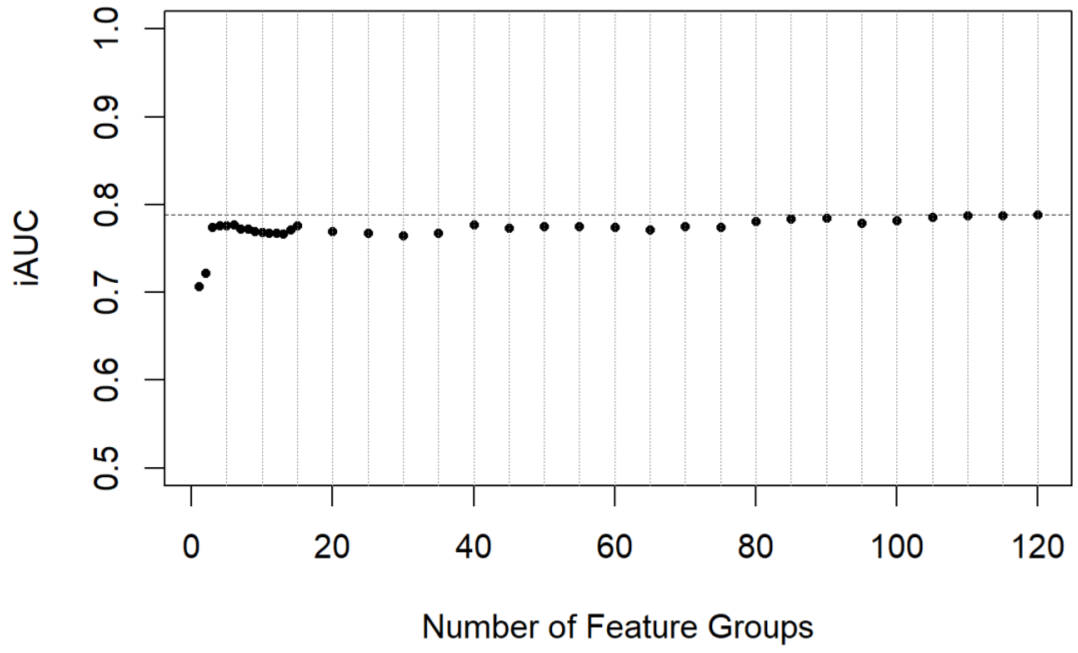


(b) Complete remission

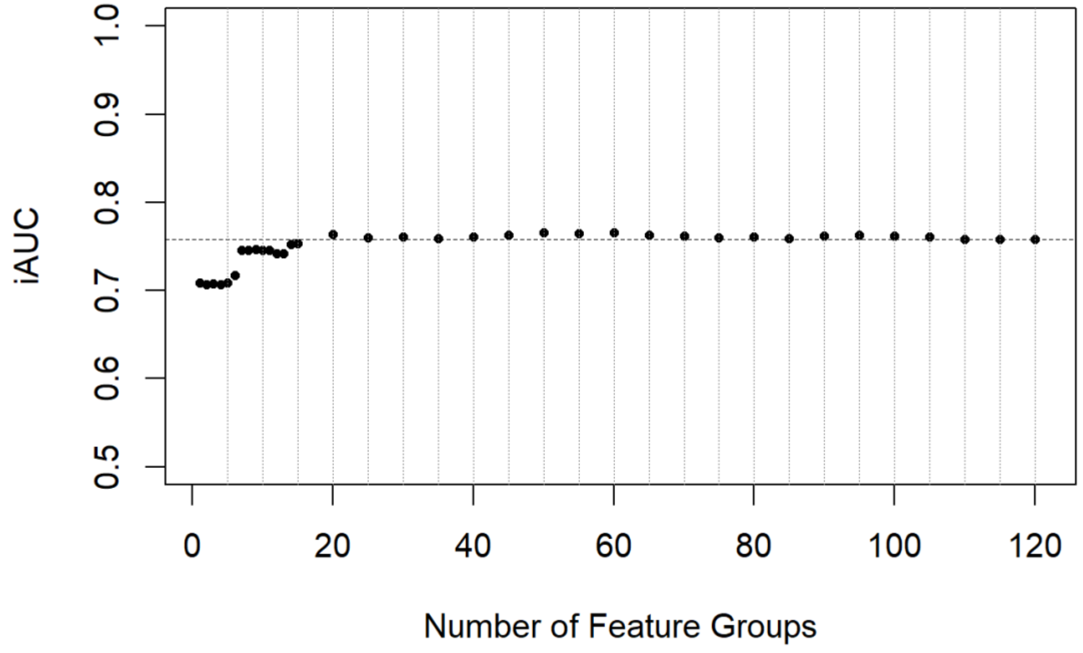

**Supplementary Figure 4** The 4 additional features which help us understand the driving factors for the predictive features.

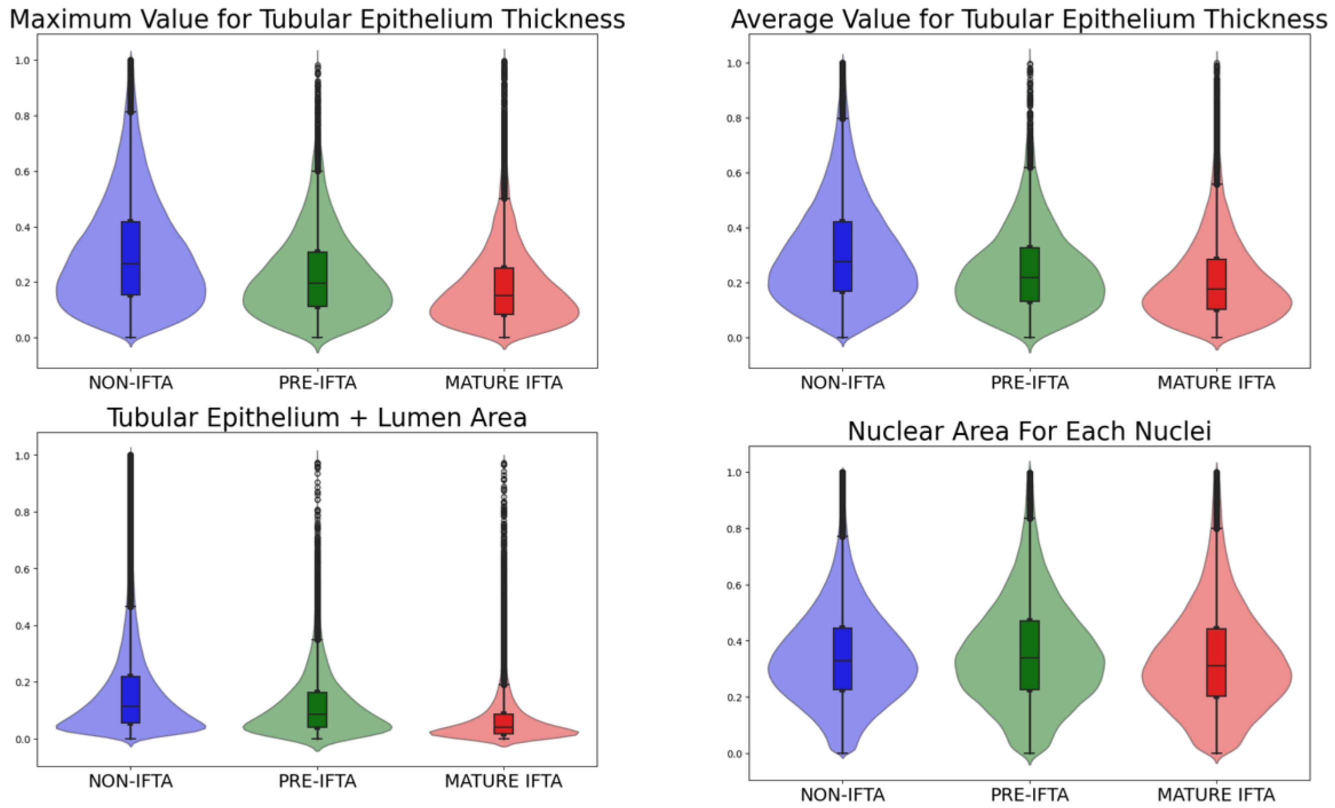

**Supplementary Figure 5** Detailed visualization of feature extraction for **a.** thickness calculation, **b.** diameter, **c.** smoothness and **d.** nuclear location features

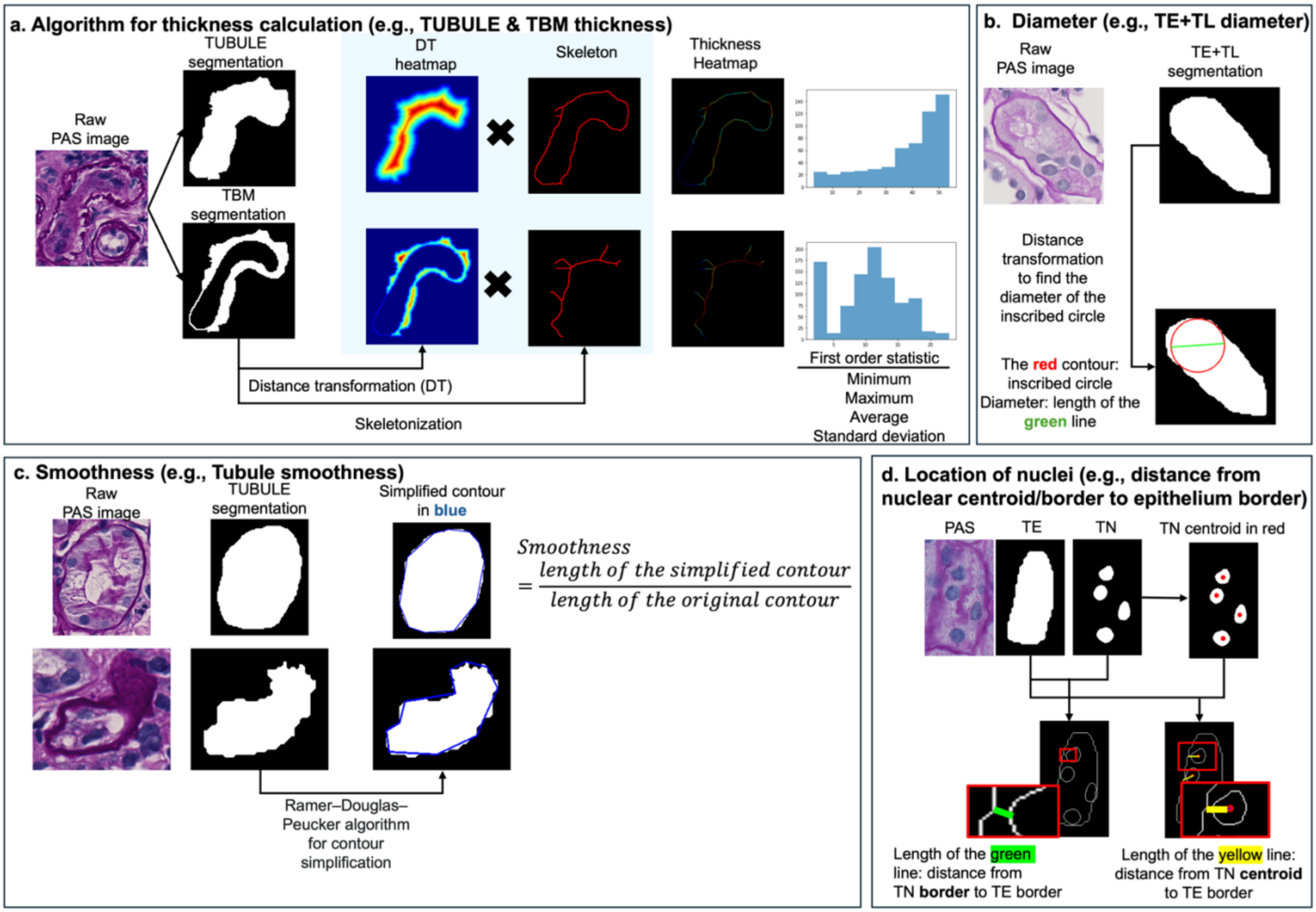

**Supplementary Table 1:** Feature names and descriptions of 99 tubule-level features

| Feature name | Description of feature |
|---|---|
| TBM_AREA | area of tubular basement membranes |
| TE_LMEN_AREA | area of tubular epithelium + tubular lumen[1] |
| TUBULE_AREA | area of TUBULE[2] (tubular epithelium, lumen, and basement membrane) |
| LMEN_AREA | area of tubular lumen |
| TE_AREA | area of tubular epithelium |
| NCLEI_AREA | area of tubular nuclei |
| TUBULE_DAMT | diameter of TUBULE (tubular epithelium, lumen, and basement membrane) |
| LMEN_DAMT | diameter of tubular lumen |
| TE_LMEN_DAMT | diameter of tubular epithelium and tubular lumen |
| TBM_THICK_MAX | maximum thickness of tubular basement membranes |
| TBM_THICK_MIN | minimum thickness of tubular basement membranes |
| TBM_THICK_AVE | average thickness of tubular basement membranes |
| TBM_THICK_STD | standard deviation of the thickness of tubular basement membranes |
| TE_THICK_MAX | maximum thickness of tubular epithelium |
| TE_THICK_MIN | minimum thickness of tubular epithelium |
| TE_THICK_AVE | average thickness of tubular epithelium |
| TE_THICK_STD | standard deviation of the thickness of tubular epithelium |
| TUBULE_THICK_MAX | maximum thickness of TUBULE |
| TUBULE_THICK_MIN | minimum thickness of TUBULE |
| TUBULE_THICK_AVE | average thickness of TUBULE |
| TUBULE_THICK_STD | standard deviation of the thickness of TUBULE |
| LMEN_THICK_MAX | maximum thickness of tubular lumen |
| LMEN_THICK_MIN | minimum thickness of tubular lumen |
| LMEN_THICK_AVE | average thickness of tubular lumen |
| LMEN_THICK_STD | standard deviation of the thickness of tubular lumen |
| TE_LMEN_THICK_MAX | maximum thickness of tubular epithelium |
| TE_LMEN_THICK_MIN | minimum thickness of tubular epithelium |
| TE_LMEN_THICK_AVE | average thickness of tubular epithelium |
| TE_LMEN_THICK_STD | standard deviation of the thickness of tubular epithelium |
| LMEN_SMOOTH_10 | smoothness of the border of the tubular lumen (Ramer-Douglas-Peucker epsilon = 10) |
| LMEN_SMOOTH_15 | smoothness of the border of the tubular lumen (Ramer-Douglas-Peucker epsilon = 15) |
| LMEN_SMOOTH_20 | smoothness of the border of the tubular lumen (Ramer-Douglas-Peucker epsilon = 20) |
| LMEN_SMOOTH_30 | smoothness of the border of the tubular lumen (Ramer-Douglas-Peucker epsilon = 30) |
| TBM_SMOOTH_10 | smoothness of the outer border of the tubular basement membrane (Ramer-Douglas-Peucker epsilon = 10) |
| TBM_SMOOTH_15 | smoothness of the outer border of the tubular basement membrane (Ramer-Douglas-Peucker epsilon = 15) |

| | |
|---|---|
| TBM_SMOOTH_20 | smoothness of the outer border of the tubular basement membrane (Ramer-Douglas-Peucker epsilon = 20) |
| TBM_SMOOTH_30 | smoothness of the outer border of the tubular basement membrane (Ramer-Douglas-Peucker epsilon = 30) |
| TE_LMEN_SMOOTH_10 | smoothness of the outer border of the tubular epithelium (Ramer-Douglas-Peucker epsilon = 10) |
| TE_LMEN_SMOOTH_15 | smoothness of the outer border of the tubular epithelium (Ramer-Douglas-Peucker epsilon = 15) |
| TE_LMEN_SMOOTH_20 | smoothness of the outer border of the tubular epithelium (Ramer-Douglas-Peucker epsilon = 20) |
| TE_LMEN_SMOOTH_30 | smoothness of the outer border of the tubular epithelium (Ramer-Douglas-Peucker epsilon = 30) |
| TE_LMEN_TBM_AREA_R | area ratio between tubular epithelium + tubular lumen and tubular basement membrane |
| TE_TBM_AREA_R | area ratio between tubular epithelium and tubular basement membrane |
| LMEN_TBM_AREA_R | area ratio between tubular lumen and tubular basement membrane |
| TUBULE_TBM_AREA_R | area ratio between TUBULE and tubular basement membrane |
| TE_LMEN_TUBULE_AREA_R | area ratio between tubular epithelium + tubular lumen and TUBULE |
| LMEN_TE_LMEN_AREA_R | area ratio between tubular lumen and tubular epithelium + tubular lumen |
| TE_TE_LMEN_AREA_R | area ratio between tubular epithelium and tubular epithelium + tubular lumen |
| LMEN_TUBULE_AREA_R | area ratio between tubular lumen and TUBULE |
| TE_TUBULE_AREA_R | area ratio between tubular epithelium and TUBULE |
| LMEN_TE_AREA_R | area ratio between tubular lumen and tubular epithelium |
| LMEN_DAMT_TBM_THICK_R | ratio between diameter of tubular lumen and average thickness of tubular basement membranes |
| TUBULE_DAMT_TBM_THICK_R | ratio between diameter of TUBULE and average thickness of tubular basement membranes |
| TE_LMEN_DAMT_TBM_THICK_R | ratio between diameter of tubular epithelium + tubular lumen and average thickness of tubular basement membranes |
| TE_TBM_THICK_R | ratio between average thickness of tubular epithelium and tubular basement membrane |
| LMEN_DAMT_TUBULE_DAMT_R | ratio between diameter of tubular lumen and diameter of TUBULE |
| LMEN_DAMT_TE_LMEN_DAMT | ratio between diameter of tubular lumen and diameter of tubular epithelium + tubular lumen |
| LMEN_DAMT_TE_THICK_R | ratio between diameter of tubular lumen and average thickness of tubular epithelium |
| TUBULE_DAMT_TE_LMEN_DAMT_R | ratio between diameter of TUBULE and diameter of tubular epithelium + tubular lumen |
| TE_THICK_TUBULE_DAMT_R | ratio between average thickness of tubular epithelium and diameter of TUBULE |
| TE_TE_LMEN_DAMT_THICK_R | ratio between diameter of tubular epithelium and average thickness of tubular epithelium + tubular lumen |
| TBM_DAMT_TUBULE_DAMT_R | ratio between diameter of tubular basement membranes and diameter of TUBULE |
| LMEN_DAMT_TBM_DAMT_R | ratio between diameter of tubular lumen and diameter of tubular basement membranes |
| TBM_DAMT_TE_LMEN_DAMT_R | ratio between diameter of tubular basement membranes and diameter of tubular epithelium + tubular lumen |

| | |
|---|---|
| TE_THICK_TBM_DAMT_R | ratio between average thickness of tubular epithelium and diameter of tubular basement membrane |
| TE_DAMT_TE_LMEN_DAMT_R | ratio between diameter of tubular epithelium and diameter of tubular epithelium and tubular lumen |
| TE_DAMT_TUBULE_DAMT_R | ratio between diameter of tubular epithelium and diameter of TUBULE |
| LMEN_DAMT_TE_DAMT_R | ratio between diameter of tubular lumen and diameter of tubular epithelium |
| TE_DAMT_TBM_DAMT_R | ratio between diameter of tubular epithelium and diameter of tubular basement membranes |
| TE_DAMT_TBM_THICK_R | ratio between diameter of tubular epithelium and average thickness of tubular basement membranes |
| NCLEI_TE_AREA_R | area ratio between tubular nuclei and tubular epithelium |
| NCLEI_TE_LMEN_AREA_R | area ratio between tubular nuclei and tubular epithelium + tubular lumen |
| NCLEI_TUBULE_AREA_R | area ratio between tubular nuclei and TUBULE |
| LMEN_NCLEI_AREA_R | area ratio between tubular lumen and tubular nuclei |
| NCLEI_TBM_AREA_R | area ratio between tubular nuclei and tubular basement membrane |
| NCLEI_LMEN_CENT_DIS_MAX | maximum distance between the center of the nuclei and the border of the lumen |
| NCLEI_LMEN_CENT_DIS_MIN | minimum distance between the center of the nuclei and the border of the lumen |
| NCLEI_LMEN_CENT_DIS_AVE | average distance between the center of the nuclei and the border of the lumen |
| NCLEI_LMEN_CENT_DIS_STD | standard deviation of the distance between the center of the nuclei and the border of the lumen |
| NCLEI_TBM_CENT_DIS_MAX | maximum distance between the center of the nuclei and the outer border of the tubular basement membranes |
| NCLEI_TBM_CENT_DIS_MIN | minimum distance between the center of the nuclei and the outer border of the tubular basement membranes |
| NCLEI_TBM_CENT_DIS_AVE | average distance between the center of the nuclei and the outer border of the tubular basement membranes |
| NCLEI_TBM_CENT_DIS_STD | standard deviation of the distance between the center of the nuclei and the outer border of the tubular basement membranes |
| NCLEI_LMEN_BORDER_DIS_MAX | maximum distance between the border of the nuclei and the border of the lumen |
| NCLEI_LMEN_BORDER_DIS_MIN | minimum distance between the border of the nuclei and the border of the lumen |
| NCLEI_LMEN_BORDER_DIS_AVE | average distance between the border of the nuclei and the border of the lumen |
| NCLEI_LMEN_BORDER_DIS_STD | standard deviation of the distance between the border of the border of the lumen |
| NCLEI_TBM_BORDER_DIS_MAX | maximum distance between the border of the nuclei the outer border of the tubular basement membranes |
| NCLEI_TBM_BORDER_DIS_MIN | minimum distance between the border of the nuclei the outer border of the tubular basement membranes |
| NCLEI_TBM_BORDER_DIS_AVE | average distance between the border of the nuclei and the outer border of the tubular basement membranes |
| NCLEI_TBM_BORDER_DIS_STD | standard deviation of the distance between the border of the outer border of the tubular basement membranes |

| | |
|---|---|
| NCLEI_TE_BORDER_DIS_MAX | maximum distance between the border of the nuclei the outer border of the tubular epithelium |
| NCLEI_TE_BORDER_DIS_MIN | minimum distance between the border of the nuclei the outer border of the tubular epithelium |
| NCLEI_TE_BORDER_DIS_AVE | average distance between the border of the nuclei and the outer border of the tubular epithelium |
| NCLEI_TE_BORDER_DIS_STD | standard deviation of the distance between the border of the outer border of the tubular epithelium |
| NCLEI_TE_BORDER_CENT_MAX | maximum distance between the center of the nuclei and the outer border of the tubular epithelium |
| NCLEI_TE_BORDER_CENT_MIN | minimum distance between the center of the nuclei and the outer border of the tubular epithelium |
| NCLEI_TE_BORDER_CENT_AVE | average distance between the center of the nuclei and the outer border of the tubular epithelium |
| NCLEI_TE_BORDER_CENT_STD | standard deviation of the distance between the center of the outer border of the tubular epithelium |
| 1: In the description of feature column: tubular epithelium + tubular lumen represents we consider epithelium and lumen as a complete structure | |
| 2: TUBULE represents tubular epithelium + tubular lumen + tubular basement membrane | |

**Supplementary Table 2:** Top feature groups predictive of (a) disease progression and (b) complete proteinuria remission. The representative features within each feature group are marked in bold. See **Supplementary Methods S4** for predetermined rules used in selecting a representative pathomic feature within each of the selected top feature group. Note that all top features reflected tubule-level features that were aggregated to the patient-level using summary statistics. Therefore the "mean," "standard deviation," or "skewness" prefixes in the feature name refers to the mean, standard deviation, or skewness across all tubules within a patient, respectively, whereas features with "minimum", "maximum" or "average" in the middle of the feature name refers to the minimum or average of multiple measurements taken within a tubule, respectively.

(a) Disease progression

| Feature Group | Description of feature |
|---|---|
| Feature Grouping 69 | **Mean of ratios between the area of the tubular epithelium and the area of the tubule** |
| Feature Grouping 1 | **Mean of area of the tubular basement membranes** |
| Feature Grouping 1 | Standard deviation of area of tubular basement membranes |
| Feature Grouping 102 | **Mean of minimal distance between the center of the nuclei and the border of the tubular lumen** |
| Feature Grouping 102 | Mean of minimal distance between the border of the nuclei and the border of the tubular lumen |

(b) Complete remission

| Feature Group | Description of feature |
|---|---|
| Feature Grouping 18 | Standard deviation of maximum thickness of tubular basement membranes |
| Feature Grouping 18 | **Standard deviation of average thickness of tubular basement membranes** |
| Feature Grouping 18 | Standard deviation of standard deviation of tubular basement membranes thickness |
| Feature Grouping 18 | Standard deviation of ratios of tubular basement membranes area to tubular area |
| Feature Grouping 18 | Standard deviation of ratios of tubular epithelium + lumen area to tubular area |
| Feature Grouping 18 | Standard deviation of ratios of tubular epithelium area to tubular area |
| Feature Grouping 18 | Standard deviation of ratios of tubular basement membrane thickness to tubule diameter (i.e., maximum thickness) |
| Feature Grouping 18 | Standard deviation of ratios of maximum tubular epithelium + lumen thickness to tubule diameter (i.e., maximum thickness) |
| Feature Grouping 18 | Standard deviation of ratios of maximum tubular basement membrane thickness to tubule diameter |
| Feature Grouping 18 | Standard deviation of ratios of maximum tubular basement membrane thickness to tubular epithelium + lumen diameter (i.e., maximum thickness) |
| Feature Grouping 89 | **Skewness of ratios between the area of the nuclei and the area of the tubular epithelium** |

| | |
|---|---|
| Feature Grouping 89 | Kurtosis of ratios between the area of the nuclei and the area of the tubular epithelium |
| Feature Grouping 91 | **Skewness of ratios between the area of the nuclei and the area of the tubular epithelium + lumen** |
| Feature Grouping 91 | Kurtosis of ratios between the area of the nuclei and the area of the tubular epithelium + lumen |
| Feature Grouping 19 | **Skewness of maximum thickness of tubular basement membranes** |
| Feature Grouping 19 | Kurtosis of maximum thickness of tubular basement membranes |
| Feature Grouping 19 | Skewness of standard deviation of tubular basement membranes thickness |
| Feature Grouping 19 | Kurtosis of standard deviation of tubular basement membranes thickness |
| Feature Grouping 57 | **Skewness of smoothness of the outer border of the tubular epithelium** |
| Feature Grouping 57 | Kurtosis of smoothness of the outer border of the tubular epithelium |
| Feature Grouping 98 | **Standard deviation of ratio between the area of the nuclei and the area of the tubular basement membrane** |
| Feature Grouping 102 | **Mean of minimal distance between the center of the nuclei and the border of the tubular lumen** |
| Feature Grouping 102 | Mean of minimal distance between the border of the nuclei and the border of the tubular lumen |
